# Transparent Implementation and Benchmarking of ACMG/AMP Variant Classification in fastVEP

**DOI:** 10.64898/2026.11.16.26368228

**Authors:** Kuan-lin Huang, Mythily Ganapathi, Wendy Chung

**Author notes:** Corresponding author: Kuan-lin Huang. Co-first author (Kuan-lin Huang, Mythily Ganapathi).

## Abstract

**Background:** The ACMG/AMP framework is the standard for germline sequence variant interpretation, but applying it in scalable automated software requires operational choices that the original framework and later ClinGen guidance do not always specify. We implemented an ACMG/AMP classification engine within fastVEP and evaluated which implementation choices improved agreement with expert-curated classifications without increasing concerning opposite-direction calls.

**Methods:** The classifier implements all 28 ACMG/AMP criteria and can evaluate 15 from a sites-only VCF using nine public evidence sources. For selecting the optimal classifier, we compared symmetric and hybrid evidence-combining schemes, pathogenic class cutoffs, population-frequency thresholds, and benign-evidence thresholds against 673,660 ClinVar variants with review status of two stars or higher from April 2026. Clinical geneticists reviewed opposite-direction calls. We then applied the selected final classifier unchanged to 116,078 variants that newly reached the two-star threshold in the October 2026 ClinVar release.

**Results:** The final fastVEP ACMG/AMP classifier uses the Tavtigian point scale on the pathogenic side and the 2015 ACMG/AMP combining table on the benign side. Using the 2015 table on both sides reduced pathogenic-side recall from 62.5 to 57.1 percent and lost 4,900 same-direction calls while avoiding 10 opposite-direction pathogenic calls. Using points on both sides increased benign-side recall from 66.4 to 77.0 percent, but increased opposite-direction benign calls from 27 to 399 and reduced recall of ClinVar uncertain variants from 97.1 to 69.5 percent. The 6-point Likely pathogenic cutoff was retained because lowering it to 5 points increased pathogenic-side recall to 73.9 percent but also classified 6,003 additional ClinVar uncertain variants and 17 additional ClinVar benign-side variants as pathogenic-side. The final classifier reproduced ClinVar’s exact five-tier class for 431,727 variants or 64.1 percent and its pathogenic, benign, or uncertain direction for 534,335 variants or 79.3 percent. Only 51 calls or 0.0076 percent were opposite in direction to ClinVar. On the later variants, same-direction agreement was 72.6 percent and 20 calls were opposite in direction.

**Conclusions:** The rules used to combine evidence affected classification more than several individual numeric thresholds. The hybrid scheme increased pathogenic-side recall while preventing a single Supporting benign criterion from producing a directional call. This conservative operating profile is suited to large-scale variant triage because it makes high-confidence calls while leaving variants uncertain when the available evidence does not support expert-level resolution. fastVEP is a research and decision-support tool and does not replace case-specific expert review.

## Introduction

The 2015 joint standards of the American College of Medical Genetics and Genomics and the Association for Molecular Pathology define 28 evidence criteria and a five-tier system for classifying germline sequence variants as pathogenic, likely pathogenic, uncertain significance, likely benign, or benign (Richards et al. 2015). The framework created a common vocabulary for diagnostic genetics, but laboratories can still reach different classifications because evidence strength, quantitative thresholds, and the treatment of conflicting evidence relies on judgment (Harrison et al. 2017).

ClinGen has addressed many of these gaps through general recommendations from the Sequence Variant Interpretation Working Group and gene-specific specifications from Variant Curation Expert Panels (Rehm et al. 2015).

Later recommendations grade PVS1 by predicted loss-of-function consequence, calibrate PP3 and BP4 by predictor strength, reduce PM2 to supporting strength, and define dedicated rules for splice evidence (Abou Tayoun et al. 2018; ClinGen SVI 2020; Pejaver et al. 2022; Walker et al. 2023). The Bayesian interpretation of the framework also produced a point system whose class boundaries do not reproduce every result of the original combining table (Tavtigian et al. 2018, 2020). Two analysts can therefore apply the same criteria but assign different classes because they combine the evidence differently.

The central problem for large-scale variant classification is not simply to maximize agreement with a reference database. A useful classifier should approximate expert geneticist judgment by making directional calls when the available evidence justifies them and by preserving uncertainty when there is insufficient phenotype, phase, segregation, or functional data to definitively classify the variant.

fastVEP is an open-source Rust reimplementation of the Ensembl Variant Effect Predictor that combines consequence prediction with direct access to clinical, population, conservation, and pathogenicity annotations (Huang 2026; McLaren et al. 2016). Its consequence annotation, compatibility, and throughput are described separately (Huang 2026). The present study evaluates the ACMG/AMP classification layer within the same annotation pass.

We compared alternative ways to combine pathogenic and benign evidence, measured the effect of class cutoffs and quantitative thresholds, and examined the classifications that remained opposite to ClinVar (e.g., pathogenic or likely pathogenic being called as benign or likely benign, or vice versa) after calibration. The operating objective was to approximate expert interpretation at scale while minimizing opposite calls and avoiding pathogenic or benign calls based on weak isolated evidence. ClinVar classifications with review status of two stars or higher provided a large source of community-curated variants, and our expert review was used to assess the opposite-direction calls rather than assume ClinVar as the truth set.

## Methods

### fastVEP Classification Strategy

The analyses reported here used the ACMG/AMP engine in fastVEP release candidate v0.5.0-rc.1 on GRCh38. Repeating the primary 673,660-variant run with the final v0.5.0 tag produced the same variant classifications and concordance matrix, with 512 fewer firings of PM2 at supporting strength, all in mitochondrial variants. The 41 single-parameter calibration runs were not repeated with the final tag.

The engine implements all 28 ACMG/AMP criteria. PP5 and BP6 are disabled by default in accordance with the ClinGen recommendation to discontinue reputable-source criteria (Biesecker and Harrison 2018). A sites-only VCF contains no proband genotype, pedigree, phenotype, segregation, functional assay, or case-control evidence, so only 15 criteria can be applied in the present analysis. These are PVS1, PS1, PM1, PM2, PM4, PM5, PP2, PP3, BA1, BS1, BS2, BP1, BP3, BP4, and BP7. The remaining criteria require patient-level or experimental information that was not available.

fastVEP performs consequence prediction and HGVS generation before classification and reports the call associated with the transcript selected by its VEP-compatible pick order. The classifier reads public evidence from gnomAD, ClinVar, ClinGen, REVEL, SpliceAI, PhyloP, RepeatMasker, gnomAD gene constraint, and Ensembl gene models (Ioannidis et al. 2016; Jaganathan et al. 2019; Karczewski et al. 2020; Pollard et al. 2010). Population-frequency criteria use the gnomAD v4.1 exome release with ancestry-specific filtering allele frequency and quality flags (Whiffin et al. 2017; ClinGen VCEP Review Committee 2024). The classifier excludes frequency evidence in 29 highly homologous genes and evaluates curated functional evidence when such a file is provided, although no functional-evidence file was used here (Mandelker et al. 2016; Brnich et al. 2019).

Criteria that summarize related evidence are reconciled before the final class is assigned. For example, PP3 is set aside beside PS1 or PM5 for the same missense effect, and the carrier-count arm of BS2 is set aside when BS1 or BA1 has already summarized the same gnomAD observation.

The selected pathogenic-side combiner uses the point scale of Tavtigian et al. (2020). Supporting, moderate, strong, and very strong evidence contribute 1, 2, 4, and 8 points, respectively. Six to nine points produce a likely pathogenic call, and 10 or more produce a pathogenic call. The selected benign-side combiner retains the 2015 ACMG/AMP table. A stand-alone criterion or two strong criteria produce a benign call, while one strong and one supporting criterion or two supporting criteria produce a likely benign call. A call is held at uncertain significance when both directions reach a call or when strong opposing evidence is present without reaching a call.

### Comparison of Combining Rules and Thresholds

We compared three combining strategies using the evidence recorded for every variant. The strategies were the Tavtigian point scale on both sides, the 2015 ACMG/AMP table on both sides, and the selected hybrid of points on the pathogenic side with the 2015 table on the benign side. The 2015-table alternative was also run through the classifier, and replay of the selected strategy reproduced 673,641 of 673,660 calls. For the remaining 19 variants, the replay differs from the real run because the real fastVEP classifier does not trust gnomAD frequencies from unreliable sites vetoed by gnomAD’s own FILTER column.

We remeasured thresholds that are not fixed by guidance with one setting changed at a time. The analysis included 41 full-benchmark runs spanning PM2 thresholds for recessive-only and other genes, the BS1 threshold, the BS2 homozygote threshold, BP7 intronic distance, the BP7 PhyloP threshold, and the count at which BP1 is withheld in a gene with established pathogenic missense variation. Additional runs varied the BS2 carrier threshold, its allele-count floor, and the 6-point likely pathogenic and 10-point pathogenic cutoffs. For each step we recorded the additional same-direction calls and the additional opposite-direction calls. Where a curve had a clear knee, we favored the point beyond which the gain per additional opposite-direction call fell sharply. Where no knee was present, published Variant Curation Expert Panel values and a preference for avoiding unsupported directional calls informed the selected operating point.

### Evidence Sources and Inheritance

Computational evidence used REVEL v1.3 for missense variants and SpliceAI plus PhyloP for splicing-related criteria. ClinVar allele-level records supplied low-penetrance and population-frequency gates, while a ClinVar protein and splice index supported PS1, PM1, PM5, BP1, and PVS1 critical regions (Landrum et al. 2018). ClinGen gene-disease validity and curated disease attributes supplied gene-level validity, mechanism, and inheritance information (Strande et al. 2017; ClinGen 2024, 2026). Autosomal dominant and autosomal recessive tags were read directly, semidominant was treated as both, and unknown, mitochondrial, and X-linked tags were not assigned to a dominant or recessive category. Inheritance determined which PM2 and BS2 thresholds were applied.

### Reference Sets and Expert Review

The development reference was the ClinVar GRCh38 VCF dated 4 April 2026. We retained single-nucleotide variants and indels of 100 bases or fewer with review status of at least two stars. The final set contained 673,660 records, including 79,823 pathogenic, 13,989 likely pathogenic, 295,298 uncertain significance, 128,038 likely benign, and 156,512 benign records. Of these, 652,400 had two stars, 21,237 had three stars, and 23 had four stars.

ClinVar labels that combine pathogenic with likely pathogenic or benign with likely benign were assigned to the first class for exact five-tier comparisons. We emphasized agreement in direction, which groups pathogenic with likely pathogenic and benign with likely benign while retaining uncertain significance as a separate state. ClinVar was treated as a community reference rather than an assumption that every classification was correct.

For temporal evaluation, we used the ClinVar release of 4 October 2026. It contained 781,135 eligible two-star-or-higher records, including 116,078 variants that were not in the April benchmark set. Most of these variants already existed in ClinVar at lower review status in April, so this is a holdout of classifications and variants rather than an independent holdout of genes.

A clinical molecular geneticist reviewed all 51 calls that were opposite in direction to ClinVar: 48 in round 6 and the remaining 3 on 10 November 2026. The review tables included the ClinVar record, fastVEP criteria, predictor scores, population frequencies and counts, gene constraint, and ClinGen disease annotations. The reviewer saw both classifications and was an author and contributor to the project, so this review was not blinded and should be interpreted as one expert’s adjudication rather than an independent gold standard. We mapped the reviewer’s verdicts for all reviewed variants to ClinVar-supported, fastVEP-supported, neither classification supported, more evidence needed, or dependent on the disease context.

### Outcomes and Statistical Analysis

An exact call matched ClinVar’s five-tier class. A same-direction call placed the variant on the same pathogenic side, benign side, or uncertain state. An opposite-direction call placed a ClinVar pathogenic-side variant on the benign side or a ClinVar benign-side variant on the pathogenic side. These were termed reference-discordant benign and reference-discordant pathogenic calls, respectively, without assuming that ClinVar was necessarily correct.

Rates use all 673,660 April records as the denominator. The 382 records with no alternate allele received no call and were counted as nonconcordant. Confidence intervals for primary rates and the temporal rate ratio were estimated by resampling genes so that variants in the same gene remained clustered. The stored tables, figures, and manuscript numbers were regenerated from scored run outputs and checked by automated scripts.

## Results

### Hybrid Combining Rules Balanced Pathogenic Recall and Benign Error Control

The final scheme released as v38 uses the Tavtigian point scale on the pathogenic side and the 2015 ACMG/AMP table on the benign side. The two halves address different limitations of symmetric scoring. On the pathogenic side, a lone very strong criterion contributes 8 points and can reach likely pathogenic, whereas the original 2015 table has no corresponding row. On the benign side, the 2015 table requires at least two supporting criteria for likely benign and therefore prevents a single weak criterion from determining direction.

Using the 2015 table on both sides produced 529,435 same-direction calls, 4,900 fewer than the hybrid scheme, and reduced pathogenic-side recall from 62.5 to 57.1 percent. It avoided 10 reference-discordant pathogenic calls, reducing them from 24 to 14. This difference corresponded to 490 additional same-direction calls for each additional reference-discordant pathogenic call under the pathogenic point scale.

The benign-side comparison was more decisive. Using points on both sides with likely benign beginning at minus 1 point increased benign-side recall from 66.4 to 77.0 percent, but reference-discordant benign calls increased from 27 to 399. Recall of ClinVar uncertain variants fell from 97.1 to 69.5 percent because a single supporting benign criterion could move many variants from uncertain significance to likely benign. Tightening the point floor to minus 2 or minus 3 reduced both recall and opposite-direction calls, but neither setting performed as well as the 2015 table on both measures.

### Combining Rules Determined the Effect of Individual Thresholds

The PM2 threshold illustrates why a numeric cutoff cannot be interpreted separately from the rule that combines the evidence. Across strict absence and thresholds up to 5 × 10^−^□, pathogenic-side recall changed by only 0.5 percentage points under the selected pathogenic point scale. Under the 2015 table on both sides, the same sweep changed recall by 6.1 points.

The difference arose because the point scale calls a lone very strong PVS1 criterion likely pathogenic at 8 points, while the 2015 table requires PM2 to accompany PVS1 for that result. Under the table, 46,207 of 51,703 pathogenic-side calls that depended on PM2 contained only PVS1 and PM2. Under the point scale, only 5,496 calls depended on PM2 and none had that two-criterion pattern. The combining rule therefore determined how many variants the PM2 threshold could move.

Benign-side thresholds produced distinct tradeoff curves (Figure 1). BP7 intronic distance showed a clear knee at 300 nucleotides. The BS1 curve showed a knee near 1 × 10^−3^, but the selected 5 × 10^−3^ setting remained on the conservative side because lowering the threshold added opposite-direction benign calls. The BS2 homozygote sweep had no clear knee, while the dominant-gene carrier threshold of 3 × 10^−^□ was the last step before lower thresholds began to add reference-discordant benign calls.

**Figure 1.**
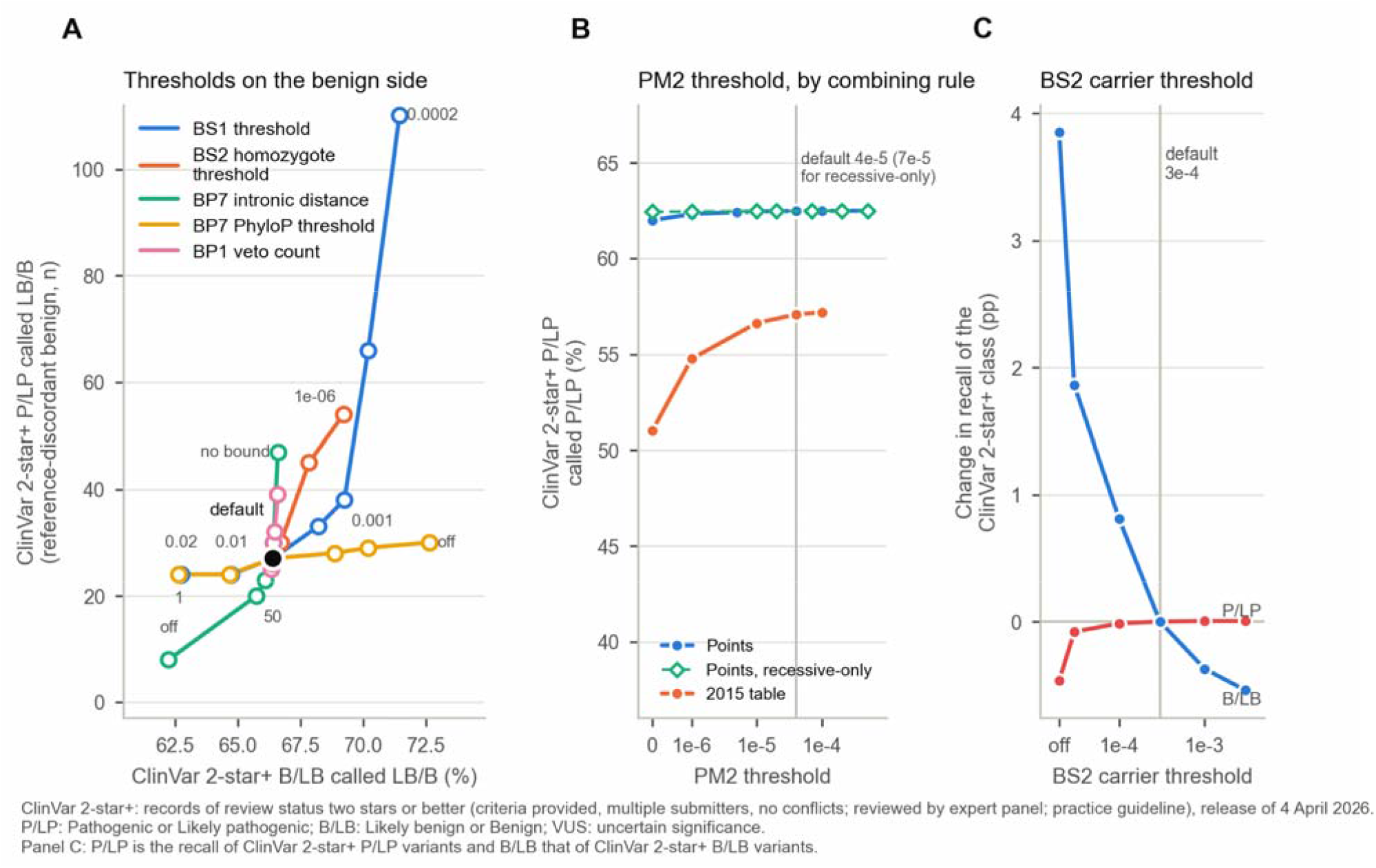
Threshold sweeps supporting selected operating points. Panel A shows the trade between correct benign-side calls and reference-discordant benign calls for five thresholds. Panel B shows that the PM2 threshold is nearly flat under the selected pathogenic point scale but consequential under the 2015 combining table. Panel C shows the BS2 carrier-threshold tradeoff after curated inheritance is applied. Filled black markers identify selected settings.

The BP7 conservation test was the strongest candidate for a future change. Removing the PhyloP requirement added 17,793 correct benign-side calls and 3 reference-discordant benign calls. The recommendation-aligned form, which also requires SpliceAI at most 0.1, added 17,170 same-direction calls for the same 3 additional reference-discordant benign calls. Because this comparison was measured on the calibration set, the final classifier retained the PhyloP threshold pending prospective evaluation.

### Class Cutoffs Dominated the Pathogenic Tradeoff

The cutoffs that define likely pathogenic and pathogenic moved more classifications than any individual numeric criterion threshold tested under the selected scheme (Figure 2). Among variants without benign evidence, 64.1 percent of variants at 5 pathogenic points were ClinVar pathogenic-side, while 35.8 percent were ClinVar uncertain. The 5-point group consisted largely of recurring combinations such as strong PVS1 plus PM2, strong PP3 plus PM2, and PM1 plus PM2 plus PM5.

**Figure 2.**
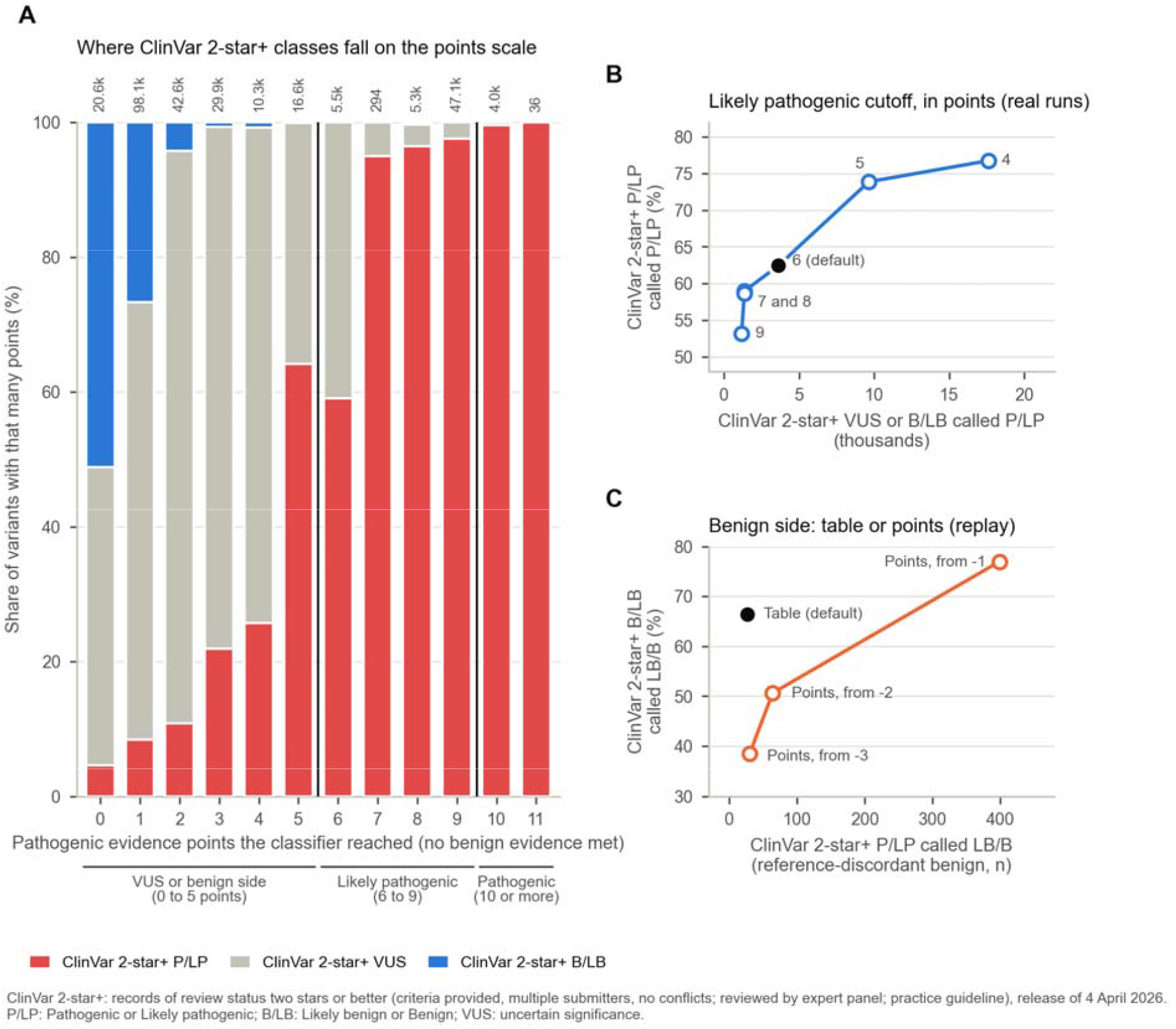
Class cutoffs dominate the pathogenic-side tradeoff. Panel A shows the ClinVar classes observed at each number of pathogenic points among variants without benign evidence. Panel B shows real runs in which the Likely pathogenic cutoff was varied. Panel C compares the 2015 benign combining table with point-based benign calls replayed from the recorded evidence.

Lowering the likely pathogenic cutoff from 6 to 5 points increased pathogenic-side recall from 62.5 to 73.9 percent and recovered 10,676 ClinVar pathogenic-side variants. It also classified 6,003 additional ClinVar uncertain variants and 17 additional ClinVar benign-side variants as pathogenic-side. Same-direction agreement increased by 4,673 calls, but exact agreement fell by 3,670 because the added calls often crossed a tier boundary not supported by ClinVar. The selected 6-point cutoff retains the published point-system calibration and avoids using a sites-only input to resolve combinations that often require phenotype, segregation, or functional evidence.

The pathogenic cutoff had little effect on direction because it separates two tiers on the same side. Moving it from 9 to 12 points changed same-direction agreement by no more than 3 calls. It substantially changed exact five-tier agreement because 47,053 of 58,201 likely pathogenic calls sat at exactly 9 points, including 46,207 variants with PVS1 plus PM2 and no other pathogenic evidence. The 10-point threshold was retained because case-level evidence that is absent from a sites-only VCF is often what distinguishes pathogenic from likely pathogenic in laboratory practice.

### The Final Classifier Favored Precision and Preserved Uncertainty

The final classifier exactly matched ClinVar for 431,727 of 673,660 variants or 64.1 percent and matched direction for 534,335 or 79.3 percent (Table 2 and Figure 3). Only 51 records were opposite in direction. The gene-cluster bootstrap 95 percent interval for the opposite-direction rate was 0.0054 to 0.0100 percent.

**Table 1.**
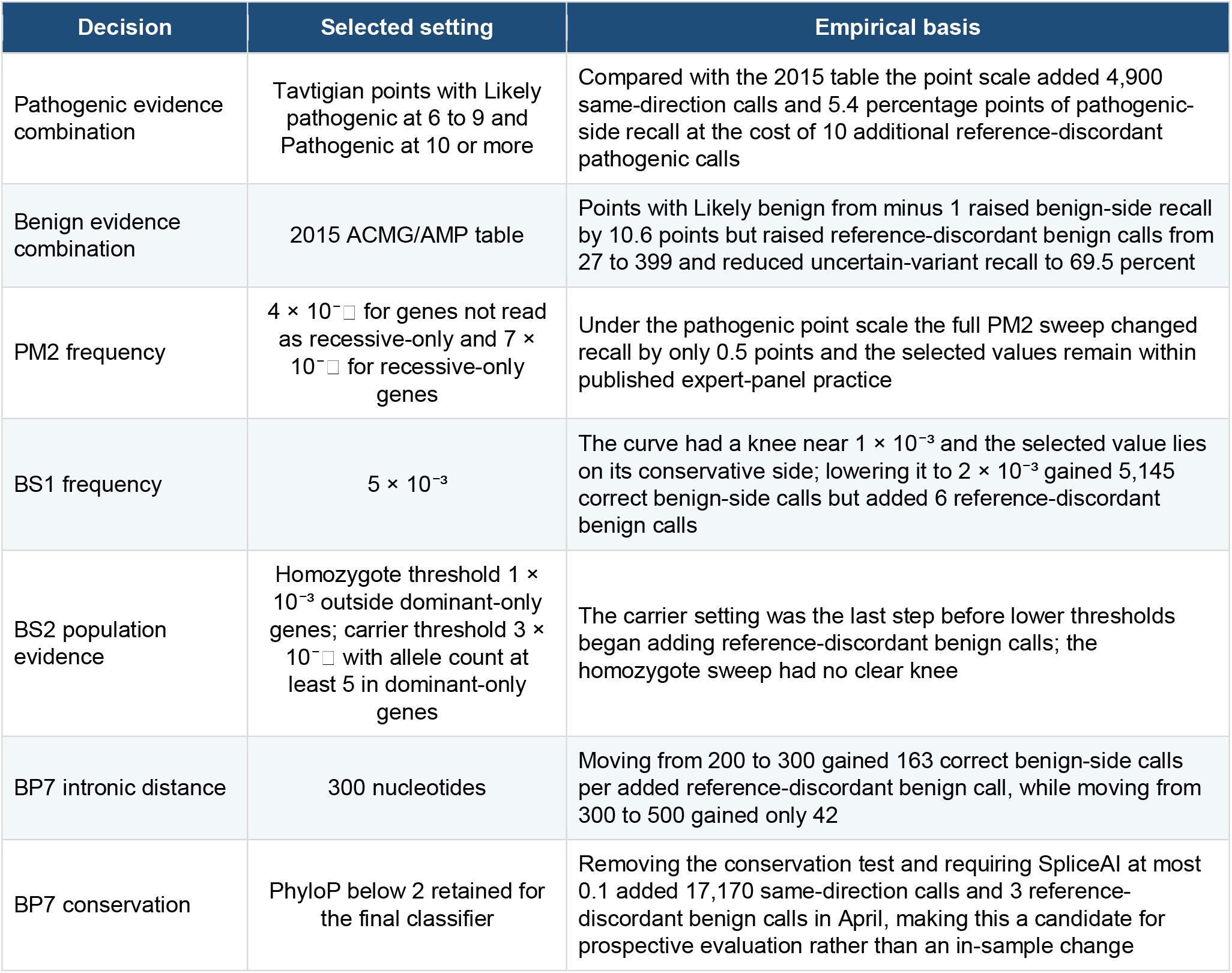
Selected combining rules and quantitative operating points.

**Table 2.**
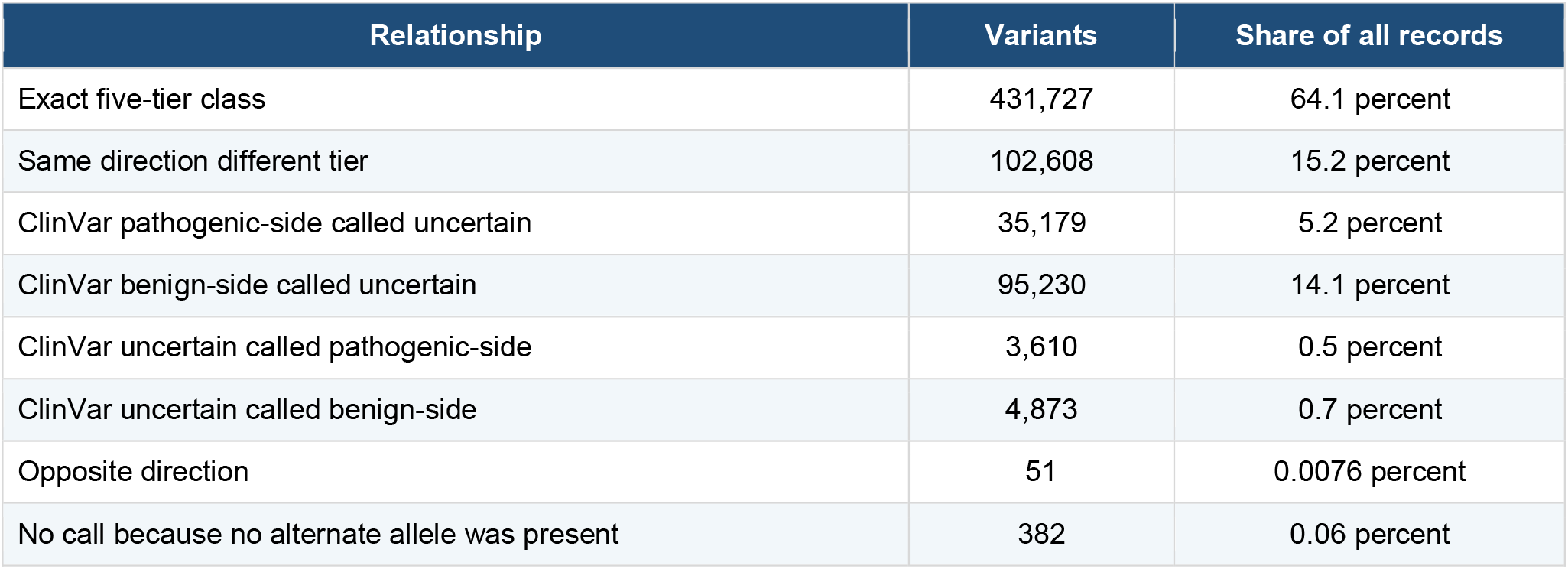
Relationship between the final classifier and ClinVar classifications.

**Figure 3.**
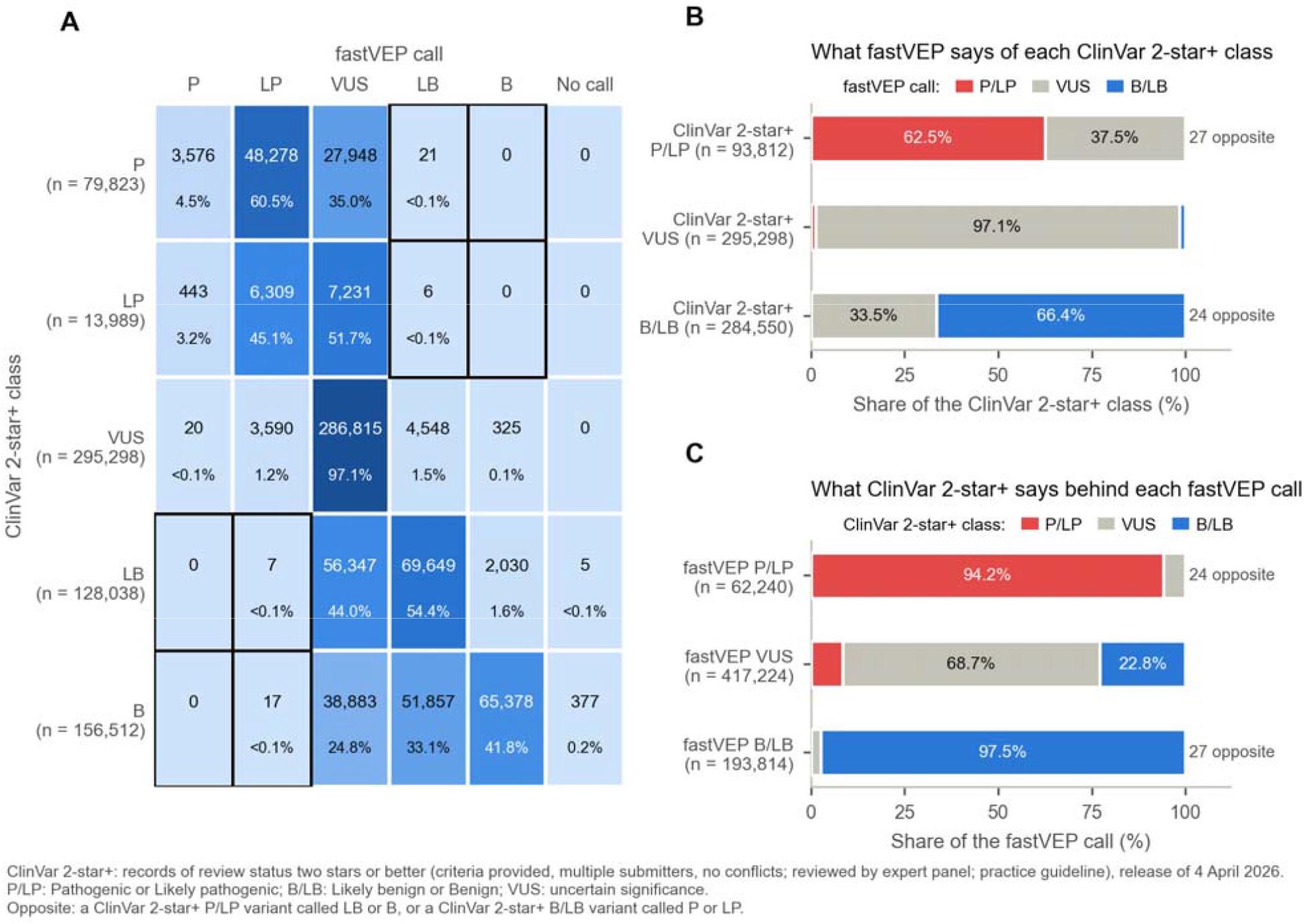
Concordance between the final classifier and ClinVar two-star-or-higher classifications. Panel A shows the five-tier confusion matrix. Panel B shows the fastVEP direction within each ClinVar direction. Panel C shows the ClinVar direction behind each fastVEP call. Outlined cells are opposite-direction calls.

Most disagreement reflected conservative classification rather than contradiction. fastVEP left 35,179 ClinVar pathogenic-side variants and 95,230 ClinVar benign-side variants at uncertain significance. Together these groups accounted for 93.6 percent of records that did not match direction. Another 8,483 ClinVar uncertain variants received a directional fastVEP call. Exact agreement was further reduced by 102,608 variants that matched direction but differed between pathogenic and likely pathogenic or between benign and likely benign.

Among 62,240 pathogenic-side fastVEP calls, 58,606 or 94.2 percent were pathogenic-side in ClinVar and 24 were opposite in direction. Among 193,814 benign-side calls, 188,914 or 97.5 percent were benign-side in ClinVar and 27 were opposite. None of the 4,039 pathogenic or 67,733 benign calls contradicted ClinVar in the April set. All 51 opposite-direction calls occurred in the likely pathogenic or likely benign tiers. These tier-specific results are in-sample and depend on the selected operating rules.

Recall varied by consequence because a sites-only input supports some evidence types more strongly than others. fastVEP called 88.4 percent of ClinVar pathogenic-side canonical splice variants and 82.6 percent of frameshift and nonsense variants on the pathogenic side, but only 17.8 percent of pathogenic-side missense variants. On the benign side, it classified 82.3 percent of intronic, 71.2 percent of synonymous, 66.3 percent of untranslated-region, and 51.8 percent of missense variants in the matching direction.

### Expert Review Characterized the Remaining Opposite Calls

The 51 opposite-direction calls concentrated in five evidence patterns (Table 3): 21 involved PVS1 on a ClinVar benign-side variant, 22 involved BP4 and BP7 on a ClinVar pathogenic-side variant, and the remaining 8 involved PM1, PP2, and PP3; BS1; or BP1.

**Table 3.** Expert assessment of the residual opposite-direction calls.

| Evidence behind the fastVEP call | Calls | Review outcome |
|---|---|---|
| PVS1 on a ClinVar benign-side variant | 21 | ClinVar 13; fastVEP 2; neither 3; more evidence 2; disorder dependent 1 |
| BP4 and BP7 on a ClinVar pathogenic-side variant | 22 | ClinVar 11; fastVEP 2; neither 6; more evidence 3 |
| PM1 PP2 and PP3 on a ClinVar benign-side variant | 3 | ClinVar supported all 3 |
| BS1 on a ClinVar pathogenic-side variant | 3 | ClinVar 2; more evidence 1 |
| BP1 on a ClinVar pathogenic missense variant | 2 | ClinVar supported both |
| All | 51 | ClinVar 31; fastVEP 4; neither 9; more evidence 6; disorder dependent 1 |

The reviewer supported ClinVar for 31 of 51 calls (61 percent), fastVEP for 4, neither classification for 9, more evidence for 6, and disorder dependence for 1. Round 6 included 48 calls, of which 29 (60 percent) supported ClinVar. The 3 later rulings supported ClinVar for VWF, where experimental evidence showed that BP7 was incorrectly applied, and TMEM216, a founder variant with a mild phenotype for which BP4 was incorrectly applied and BS1 did not apply. GTPBP3 was read as VUS on the MANE Select transcript and coded as neither classification, although its interpretation may be disorder-dependent. Thus, opposite-direction calls are high-priority review targets, not automatic errors.

Many rulings required evidence unavailable to a sites-only classifier, including disease-specific penetrance, phase, functional splicing results, transcript choice, first-exon escape from nonsense-mediated decay, and inheritance mode. A disease-specific frequency threshold could affect about 8 of the 51 calls. The review also showed that the BP4 splice path lacked a filter for variants outside the transcript, although the number affected was not measured. These findings support automated classification as structured triage rather than final clinical interpretation.

### Performance Remained Similar on the Additional Variants in the October ClinVar Release

The October release contained 116,078 variants that were not in the April benchmark set (Table 4 and Figure 4). Same-direction agreement was 72.6 percent, compared with 79.3 percent in April. Recall was 60.5 percent for pathogenic-side variants, 60.0 percent for benign-side variants, and 97.1 percent for uncertain variants.

**Table 4.** Temporal evaluation of the final classifier.

| **Set** | **Variants** | **Same-direction agreement** | **Pathogenic-side recall** | **Benign-side recall** | **Uncertain recall** | **Opposite-direction calls** |
|---|---|---|---|---|---|---|
| April development set | 673,660 | 79.3 percent | 62.5 percent | 66.4 percent | 97.1 percent | 51 |
| October release all eligible variants | 781,135 | 78.3 percent | 62.7 percent | 65.3 percent | 97.2 percent | 68 |
| October variants not in the April set | 116,078 | 72.6 percent | 60.5 percent | 60.0 percent | 97.1 percent | 20 |

**Figure 4.**
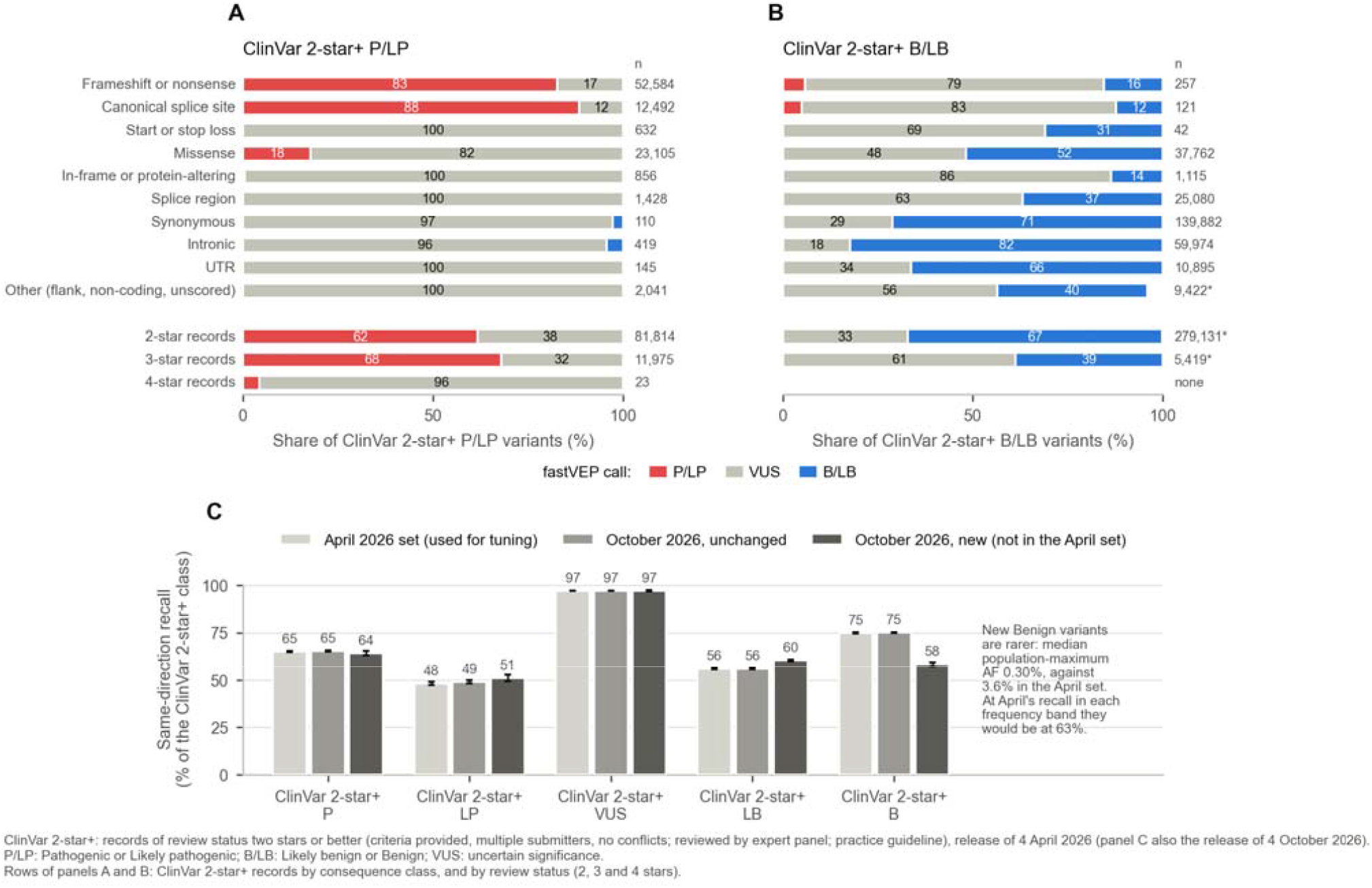
Recall by consequence review status and time. Panels A and B show fastVEP calls for ClinVar pathogenic-side and benign-side variants by consequence class and review status. Panel C compares class-specific recall in the April calibration set, October variants with an unchanged class, and October variants not in the April set.

The later set contained more benign-side variants and fewer uncertain and pathogenic-side variants. Applying April recall separately to each direction showed that class composition explained 2.9 of the 6.7 percentage-point decline, while lower within-class recall explained 3.8 points. Most of the within-class decline occurred among new benign variants, whose median maximum population frequency was 0.30 percent compared with 3.6 percent in the April set.

Twenty calls among the later variants were opposite in direction, a rate of 0.017 percent. This was 2.3 times the April rate, with a gene-cluster bootstrap 95 percent interval of 1.2 to 3.8. Ten involved PVS1 on a ClinVar benign-side variant, seven involved BP4 and BP7 on a ClinVar pathogenic-side variant, and three involved missense evidence from PP2 and PP3. The evidence patterns therefore resembled those in the April set.

The later set is not fully independent. Ninety-eight percent of its variants were already present in ClinVar in April at a lower review status, and only 3.2 percent of its genes were absent from the April benchmark. It nevertheless evaluates classifications that were not used to select the operating points or review the original opposite-direction calls.

### Discussion Principal Findings

The central result is that the evidence-combining rule determines the classifier’s operating profile. The selected hybrid uses the pathogenic point scale to recognize strong pathogenic evidence that the original table maps inefficiently, while retaining the benign table to prevent one supporting benign criterion from producing a directional call. This combination preserved more pathogenic-side recall than the table on both sides and produced far fewer opposite-direction benign calls than points on both sides.

The class cutoffs were more influential than most individual numeric thresholds. Moving the likely pathogenic cutoff from 6 to 5 points increased recall by 11.4 percentage points, but it also moved thousands of ClinVar uncertain variants to the pathogenic side. In contrast, the full PM2 sweep moved pathogenic-side recall by only 0.5 points under the selected point scale. The same PM2 threshold became consequential under the 2015 table because the table needs PM2 to accompany PVS1. Thresholds should therefore be calibrated together with the combining rule in which they operate.

The selected classifier favors precision over directional call rate. It made directional calls for 256,054 April variants, and 94.2 percent of its pathogenic-side calls and 97.5 percent of its benign-side calls matched ClinVar’s direction. Most disagreement arose because fastVEP left a ClinVar-defined variant uncertain rather than placing it on the opposite side. For a classifier that lacks phenotype, segregation, phase, and patient-level functional evidence, retaining uncertainty is consistent with the intended role of high-throughput decision support.

### Approximating Expert Judgment at Scale

The practical target of automated ACMG/AMP interpretation is not to reproduce every database label. It is to prioritize variants in a way that resembles how a geneticist weighs evidence, while making the evidence behind each call available for review. The hybrid combining scheme serves that target by treating strong pathogenic evidence differently from weak benign evidence. It allows a very strong pathogenic criterion to contribute meaningfully but requires corroboration before a weak benign observation determines direction.

The threshold optimization sweeps support conservative operating points rather than a single rule of maximum agreement. The 300-nucleotide BP7 boundary had a reproducible knee. The 5 × 10^−3^ BS1 threshold remained on the cautious side of its empirical knee, and the dominant-gene BS2 carrier threshold of 3 × 10^−^□ was the last step before lower values began adding opposite-direction benign calls. The PM2 thresholds were comparatively stable because the pathogenic point scale made many calls independent of PM2.

The BP7 conservation result shows how the framework can identify a promising future change without adopting it prematurely. The recommendation-aligned alternative gained more than 17,000 same-direction calls for 3 additional opposite-direction benign calls in April and gained 5,744 calls for 1 additional opposite-direction benign call among later variants. Those results favor prospective evaluation without the conservation requirement, but the final classifier retains the current setting because the April comparison was used during calibration.

Expert review provided a complementary measure of clinical plausibility. The reviewer supported ClinVar for 31 of 51 opposite-direction calls, supported fastVEP or neither classification in 13, requested more evidence in 6, and judged 1 to depend on the disorder. This distribution reinforces the use of opposite-direction calls as a review queue rather than as an automatic error count. It also shows why a sites-only classifier should expose its criteria and preserve VUS calls when disease-specific evidence is missing.

### Recommended Operating Configuration

For large-scale sites-only annotation, the results support the following configuration. Pathogenic evidence is combined by the Tavtigian scale with likely pathogenic at 6 points and pathogenic at 10 points. Benign evidence is combined by the 2015 ACMG/AMP table rather than a point floor that would allow one supporting criterion to call likely benign. PM2 uses 4 × 10^−^□ outside recessive-only genes and 7 × 10^−^□ in recessive-only genes, while BS1 uses 5 × 10^−3^. BS2 uses a homozygote threshold of 1 × 10^−3^ outside dominant-only genes and a carrier threshold of 3 × 10^−^□ with at least 5 observed alleles in dominant-only genes. BP7 extends to 300 intronic nucleotides, with the conservation requirement retained pending prospective validation of the recommendation-aligned alternative.

These defaults should not be interpreted as universal disease-specific thresholds. Variant Curation Expert Panel specifications should take precedence when a gene-disease pair has an applicable calibrated value. The global defaults provide a reproducible fallback for large-scale triage when disease-specific prevalence and penetrance are unavailable.

### Interpreting ClinVar Agreement

ClinVar remains the most useful large public reference for this analysis, but agreement with it is not equivalent to clinical validity. The expert reviewer did not support ClinVar for every opposite-direction call, and ClinVar submitters can use the same population frequencies, computational predictors, or previously classified variants that fastVEP reads. The reported agreement therefore reflects both genuine alignment with expert practice and partial dependence on shared evidence.

The 8,483 directional fastVEP calls on ClinVar uncertain variants are also not necessarily errors. Some ClinVar uncertain records reflect genuinely conflicting or insufficient evidence, while others may not have been fully evaluated. The six-month follow-up was too short to determine whether ClinVar would move toward or away from these calls.

Exact five-tier agreement is strongly influenced by tier boundaries. More than 100,000 variants matched direction but differed between pathogenic and likely pathogenic or between benign and likely benign. Exact agreement therefore measures both evidence direction and how the classifier handles the final tier boundary. For large-scale triage, direction, call precision, uncertain-call retention, and opposite-direction events are more informative when reported together.

### Limitations

Every threshold was selected or reviewed against the April ClinVar set. The April results are therefore in-sample, and the low opposite-direction rate reflects the operating choices selected on those variants. The October analysis provides a partial holdout, but most variants already existed in ClinVar at lower review status and almost all genes were represented in April.

One clinical molecular geneticist performed the systematic review. The reviewer was not blinded, saw only opposite-direction calls, contributed evidence to the project, and returned marked workbooks for three rounds. The categories used here were coded from those records and were not independently confirmed.

ClinVar’s two-star-or-higher subset excludes many single-submitter and conflicting records. The benchmark is limited to single-nucleotide variants and short indels and evaluates one selected transcript per variant. The same variant can have a different consequence or classification on another clinically relevant transcript.

The classifier can evaluate only 15 of 28 criteria from the sites-only input used here. It cannot observe phenotype specificity, segregation, de novo status, phase, patient-level functional evidence, or case-control enrichment. Its uncertain classifications are therefore not equivalent to the conclusions of a laboratory that has access to those data.

We did not compare fastVEP directly with another automated ACMG/AMP classifier. The study evaluates operating choices within one system and should not be interpreted as a comparative ranking of tools.

### Priorities for Prospective Evaluation

The most immediate prospective comparison is BP7 without the conservation requirement and with SpliceAI at most 0.1, as described in the splicing recommendation. The April and October results suggest a favorable exchange of additional benign-side calls for a small number of opposite-direction calls, but an independent future release is needed before changing the default.

Disease-specific population thresholds are the second priority. The global BS1, BS2, and PM2 settings cannot represent the prevalence, penetrance, and allelic contribution of every disorder. Expanding validated Variant Curation Expert Panel thresholds and associating them with the relevant gene-disease pair should improve approximation of expert judgment.

Future work should also test the selected operating profile on patient-level cases that include phenotype, segregation, phase, and functional evidence. That evaluation would measure whether variants left uncertain by the sites-only classifier are resolved in the same direction as a medical geneticist who has access to the full clinical record.

## Conclusions

Implementing ACMG/AMP classification in fastVEP required repeated translation of narrative guidance into explicit conditions, tests, and evidence dependencies. Expert review and end-to-end validation found defects that aggregate concordance could not reveal. The final classifier matched ClinVar’s direction for 79.3 percent of 673,660 development variants with 51 opposite-direction calls and for 72.6 percent of 116,078 later variants with 20 opposite-direction calls.

These results support a conservative use of automated classification. The most reliable implementation is not the one that maximizes agreement by making more calls. It is the one that documents what evidence was available, prevents dependent observations from being counted twice, preserves uncertainty when patient-level evidence is absent, and makes each remaining disagreement reviewable.

## Data Availability

fastVEP is open source under the Apache License 2.0 at https://github.com/Huang-lab/fastVEP. The platform paper is available at https://www.biorxiv.org/content/10.64898/2026.04.14.718452v2. The ACMG/AMP benchmark harness, run history, analysis scripts, figures, source tables, and reproducibility checks are available in the fastVEP repository under analysis/acmg_benchmark and manuscript/acmg. The repository stores the scored concordance matrices, criterion firing counts, configurations, and provenance records for the manuscript runs. Large annotated VCFs and source databases are not included, but their public sources and versions are documented.

## Declarations

### Author contributions

Kuan-lin Huang conceptualized the study, designed and implemented the classifier, conducted the benchmark and calibration analyses, and drafted the manuscript. Mythily Ganapathi performed the clinical review of discordant calls and contributed curated variant inputs. Wendy Chung advised on the clinical interpretation of a subset of cases. All authors reviewed, edited, and approved the final manuscript.

