## Supplementary Information for "Transparent Implementation and Benchmarking of ACMG/AMP Variant Classification in fastVEP"

### Supplementary Material

**A Fast, Robust, and Tuned Implementation of the ACMG/AMP Variant Classification in fastVEP**

#### Contents

Supplementary Figures S1 to S4; Supplementary Notes 1 to 12; Supplementary Tables S1 to S9; Appendix Tables A1 to A9. The notes give detail that the main text summarises; they do not repeat it.

The supplementary and appendix tables are the data files gathered in the workbook supplementary_tables.xlsx.

The supplementary figures show how the classifier was developed and reviewed, the supplementary notes hold the detail of the process and of the sweeps that the main text summarises, and the nine supplementary tables are the tables that summarise the data best; every other table is in the appendix. All of the tables are in one workbook, manuscript/acmg/supplementary_tables.xlsx, which has a sheet of contents and a sheet for each table (S1 to S9 and A1 to A16, of which A10 to A16 are the tables of version v38, which this document does not describe). Each table is also a set of CSV or TSV files in manuscript/acmg/data/ of the repository, which the entries below name, and each of those files is written by analysis/acmg_benchmark/scripts/13_manuscript_stats.py, 11_manuscript_runs.py, 14_temporal_holdout.py or 15_clingen_complete_export.py from the scored runs.

#### Supplementary figures

In Figures S1, S3 and S4 a ClinVar class is that of a ClinVar 2-star+ record of the release of 4 April 2026, as in the main figures, and each figure says so in a footnote. Figure S2 shows no ClinVar class.

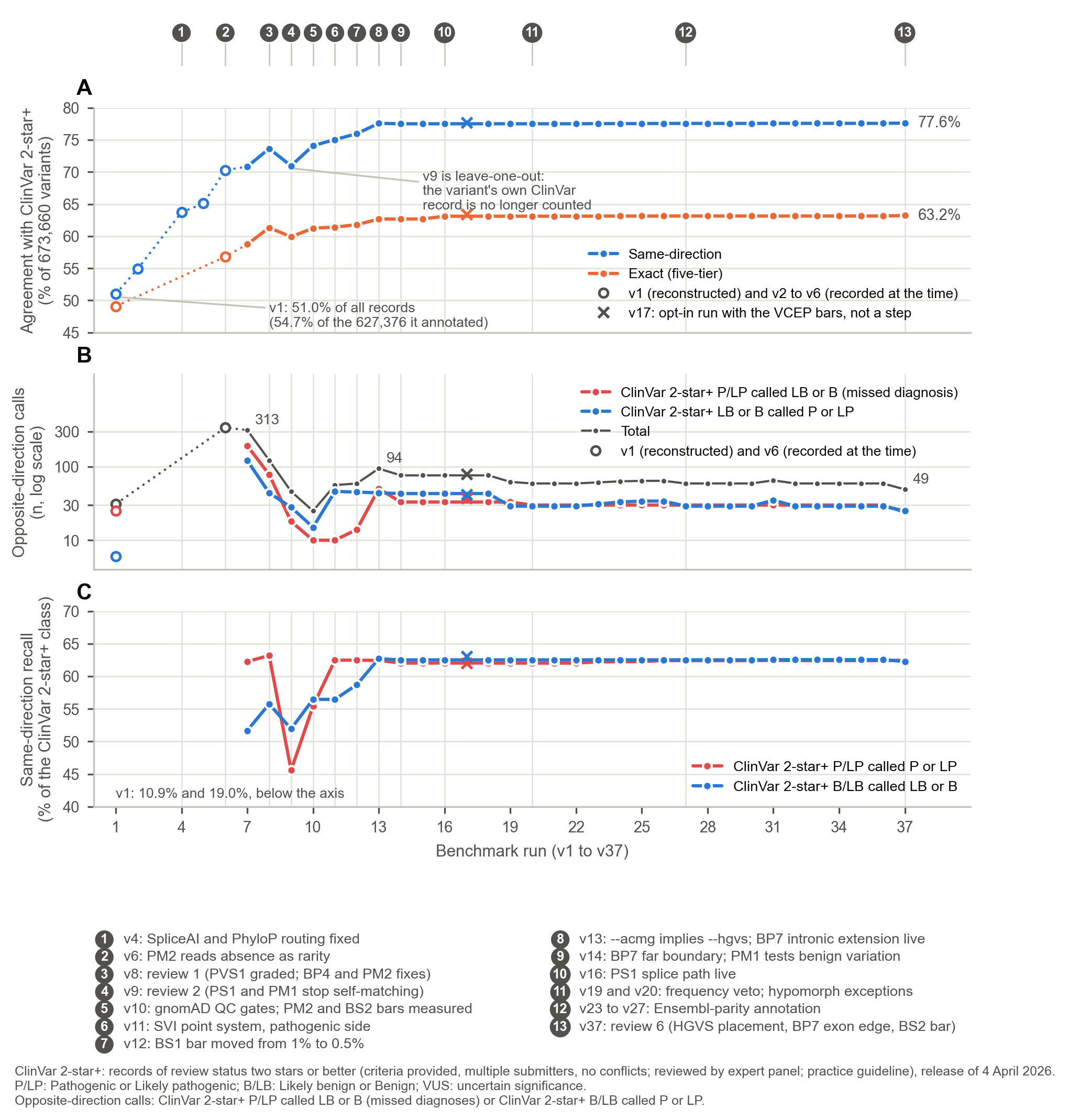

**Figure S1. Agreement with ClinVar 2-star+ classifications across the benchmark runs v1 to v37.** (A) Same-direction and exact agreement as a percentage of 673,660 variants. Open markers are v1, a matrix reconstructed from the documentation of the run, and v2 and v4 to v6, which were recorded at the time; no raw output of them was kept. v1 annotated 627,376 of the records, and its 54.7% of those is 51.0% of all records. The cross marks v17, the opt-in run with the published VCEP thresholds, which is not a step on the way and is left out of the lines. (B) Opposite-direction calls on a log scale: ClinVar 2-star+ pathogenic-side variants called benign-side (reference-discordant benign calls), ClinVar 2-star+ benign-side variants called pathogenic-side, and their total; open markers as in A, and v6 is the recorded total. (C) Same-direction recall for ClinVar 2-star+ pathogenic-side and benign-side variants; the values of v1 (10.9% and 19.0%) are below the axis. The 13 numbered markers are the changes listed under the panels. Run v3 was a partial run that was overwritten, v21 and v22 repeat v20, and v24 and v25 were intermediate runs. ClinVar’s classifications are a reference and not ground truth.

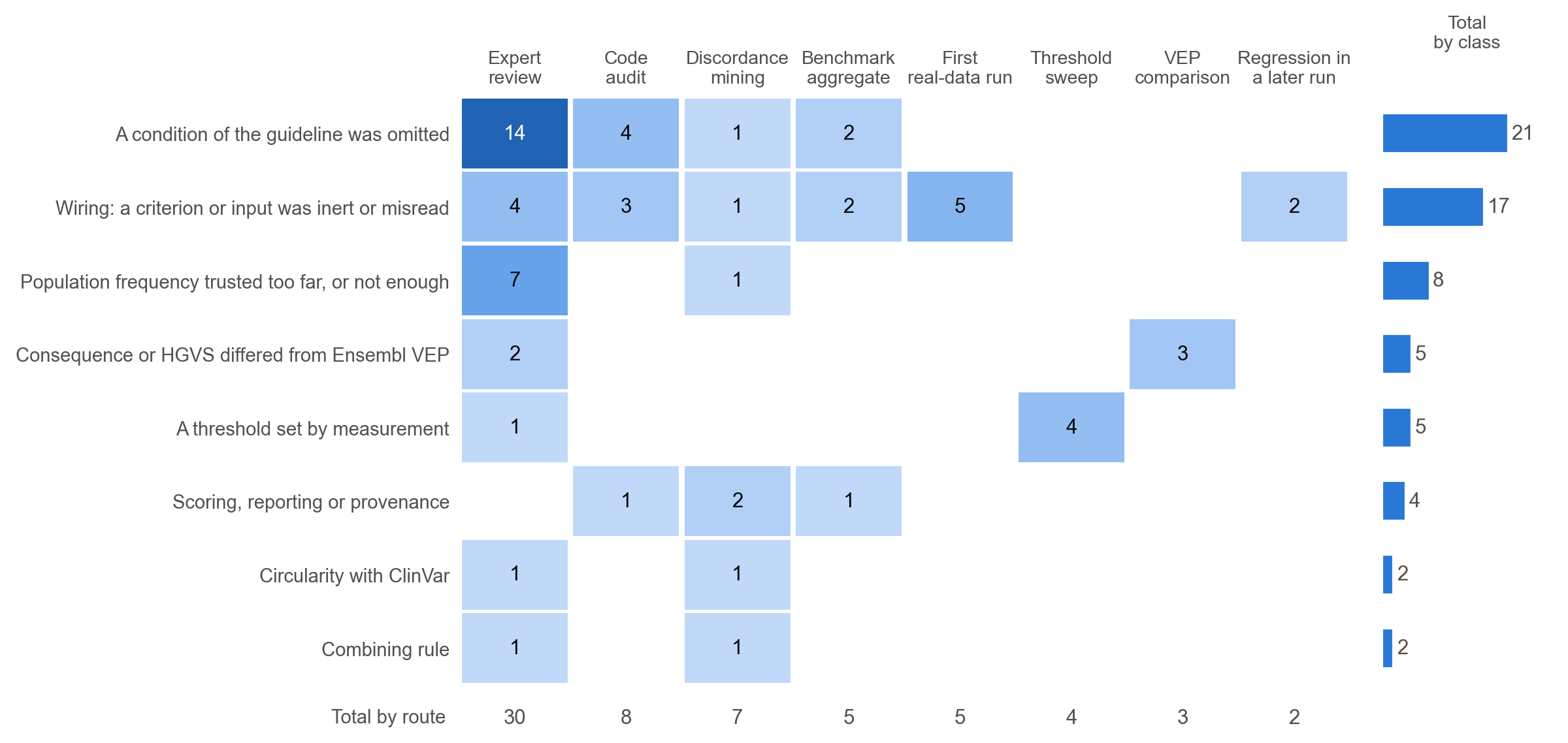

**Figure S2. The 64 recorded changes by class and by the route that first found them.** Each cell counts changes in the ledger of Supplementary Table S3. The column on the right gives the total of each class, and the row beneath the cells the total of each route. Classes and routes are our coding of the project’s record.

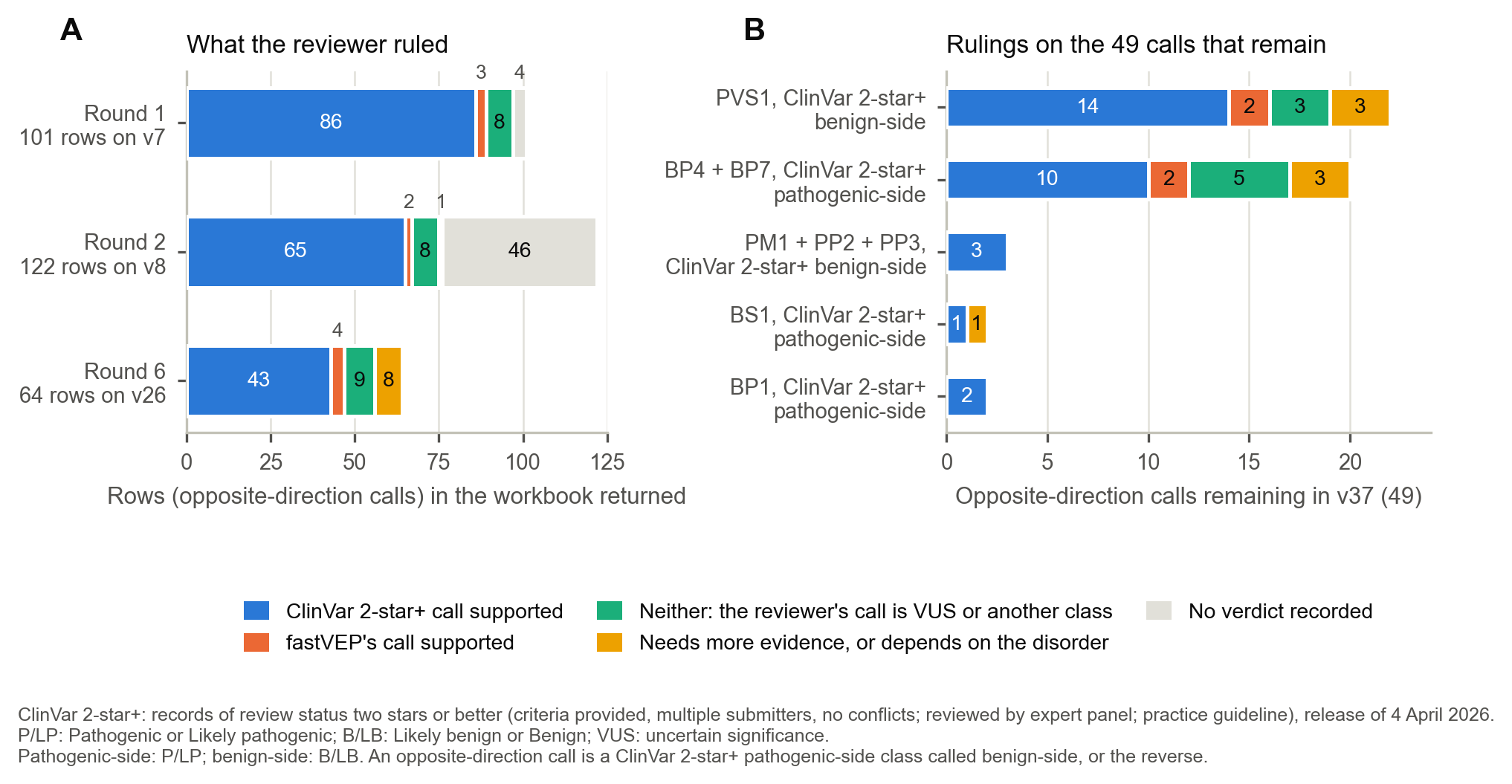

**Figure S3. What the reviewer ruled.** (A) The rulings on the rows of the three workbooks the reviewer returned, in rounds 1, 2 and 6, in five categories: the ClinVar 2-star+ call supported, fastVEP’s call supported, neither (her own call is VUS or another class), more evidence needed or dependent on the disorder, and no verdict recorded. Rounds 1 and 2 are decoded from cell colours against each workbook’s key, and round 6 is our coding of her text (Appendix Table A4). (B) The 49 opposite-direction calls of v37, by the evidence behind the call, with the rulings of the last round.

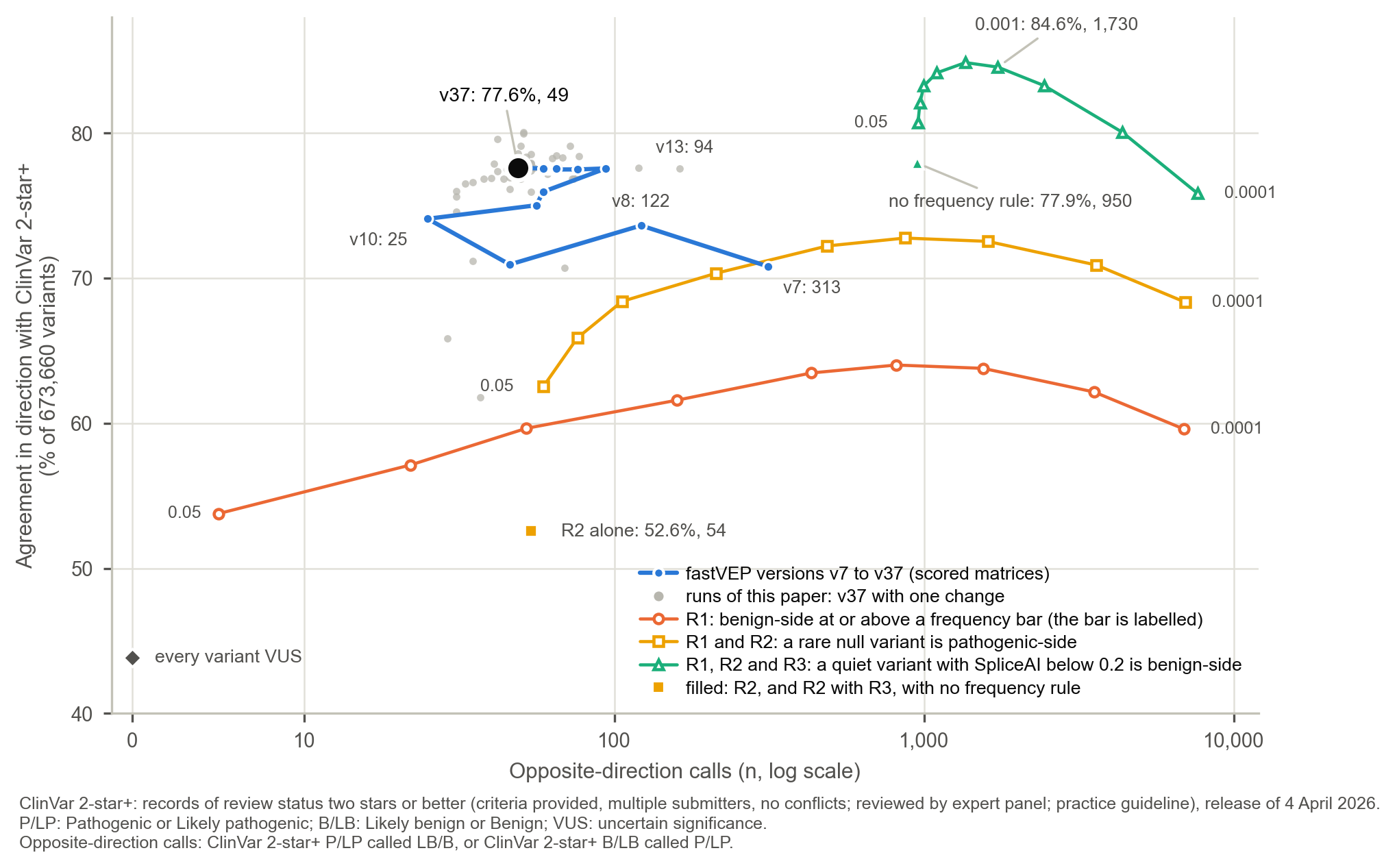

**Figure S4. Agreement in direction against opposite-direction calls.** Each point is a classifier, on 673,660 variants: the versions v7 to v37 with stored scored matrices (blue, joined in order), the runs of this paper and the earlier runs of the dominant-gene BS2 settings, each v37 with one change (grey), every variant called VUS (the diamond at zero), and three rules with no combining rule, gene-level gate or safeguard at nine frequency thresholds each, with the second rule alone and the second with the third also given with no frequency rule (the filled markers). R1 calls a variant benign-side when its population-maximum allele frequency is at or above the threshold (the thresholds at the two ends of each line are labelled, and the point at 0.001 of the last rule is annotated), R2 adds a pathogenic-side call for a stop-gained, frameshift, canonical splice or start-lost variant that is absent from gnomAD or below 1e-4, and R3 adds a benign-side call for a quiet variant (synonymous, intronic, untranslated, flanking or non-coding) with SpliceAI below 0.2. The horizontal axis is on a log scale above 10.

#### Supplementary notes

**Supplementary Note 1. The run series, and where agreement came from.**

Routes of evidence that the benchmark does not use. PS3 and BS3 are read from a curated file when one is supplied, PS2 and PM6 from the genotypes of a proband’s parents, and PM3 and BP2 from the proband’s genotype together with its other variants in the gene; none of these routes is used in the benchmark.

Runs that are not steps on the way. Run v3 was a partial run that was overwritten before it was preserved, v21 and v22 repeat v20, v24 and v25 were intermediate runs that are not versions that the project kept, and v17 is the opt-in run with the published VCEP thresholds, which v18 does not carry. We mark v17 in the figures and leave it out of every range that the text quotes. The raw output of v1 to v6 was overwritten, so v2 and v4 to v6 are quoted from the figures recorded at the time, and v1 from a matrix that was reconstructed from the documentation of the run: scored matrices are stored for 31 of the 37 runs, v7 to v37. Every rate is over the 673,660 records of the benchmark; v1 annotated only 627,376 of them, and its recorded 54.7% is over those, where over all records it is 51.0%. We could not check the denominators of v2 to v6.

Comparison with Ensembl VEP. Where a change touched the annotation layer (consequence or HGVS), every variant whose call moved was re-annotated with Ensembl VEP release 115.1 on the same GFF3 and FASTA with --pick, after confirming that both tools reported the same transcript, to establish which tool the change brought the software closer to; the written record keeps the outcome only as ledger items (3 were found by this comparison), and we do not report the comparison itself.

Where agreement came from, and where it stopped. Same-direction agreement with ClinVar rose from 51.0% in v1 (a matrix reconstructed from the run’s documentation) to 77.6% in v37 (Figure S1A; Supplementary Table S2 gives every run). Almost all of that rise came in the first 13 runs, and the largest steps were mostly plumbing and not new guidance. Agreement rose 8.8 points between v2 and v4, when SpliceAI and PhyloP results finally reached the criteria that read them (items 6 and 7 of the ledger), and 5.2 points between v5 and v6, when a variant absent from gnomAD began to count as absent for PM2 and BP4’s splice path was confined to splice questions (items 9 and 10 of the ledger), two conditions of the guidelines that the criteria had not applied. The first review (v7 to v8) added 2.8 points. The next run, v9, lost 2.7 points, and that loss was the correct result: PS1 and PM1 had been counting each variant’s own ClinVar record, and 94% of PS1’s firings were self-matches, so scoring them leave-one-out removed agreement that had never been evidence. Of the 2.7 points, about 0.6 were the cost of the review’s criteria fixes and about 2.0 were the leave-one-out scoring (RUN_VERSIONS.md). Recall of ClinVar’s pathogenic-side variants fell from 63.2% at v8 to 45.6% at v9 for the same reason, and the 62.5% of v37 is 0.7 points below the 63.2% that v8 reached by counting each variant’s own record. Agreement then rose by 3.2 points at v10, which bundled a rebuilt gnomAD database with its quality fields, the gene-level gates, and the first two measured thresholds (the PM2 threshold and the BS2 prevalence threshold), by 0.9 points each for the SVI point system (v11) and the BS1 threshold (v12), and by 1.6 points at v13, when --acmg was made to enable HGVS and the criteria that read it came alive.

After v13 the headline stopped moving. Across the next 23 runs (v14 to v37, without v17) same-direction agreement stayed between 77.50% and 77.62%, and no run moved it by more than 0.07 points. The work did not stop, and its effect shows in the other panels. Opposite-direction calls fell from 94 at v13 to 49 at v37, reference-discordant benign calls from 50 to 24, and reference-discordant pathogenic calls from 44 to 25 (Figure S1B). The path was not monotonic. Opposite-direction calls had reached a minimum of 25 at v10 and then rose to 56 at v11, when a lone PVS1 became Likely pathogenic under the point system, which gained 6,632 correct pathogenic-side calls for 31 additional reference-discordant pathogenic calls. They rose again at v13, when BP7’s intronic extension started to fire, which gained 11,339 benign-side calls for 36 additional reference-discordant benign calls. Both were trades made on purpose and recorded as trades. The matrix stored for v1 holds 31 opposite-direction calls, fewer than v37, because v1 committed to only 74,355 variants where v37 commits to 244,093; per committed call, contradictions fell from 0.042% in v1 to 0.020% in v37, with a minimum of 0.011% at v10. Recall of ClinVar’s pathogenic-side variants in direction has stayed between 62.0 and 62.5% since v11, and recall of its benign-side variants between 62.3 and 62.7% since v13 (Figure S1C). That flat line is the structural fact about the benchmark that the Results examine. The agreement figures also have an uneven past: the raw output of v1 to v6 was not kept, and v2 and v4 to v6 are quoted from the figures recorded at the time with denominators that we could not check, so their values carry less weight than the stored runs from v7.

Switching on PS1’s splice path left same-direction agreement unchanged and moved exact agreement by 2,952 calls (0.44 points), because it turned 3,793 Likely pathogenic calls into Pathogenic, a change the same-direction measure does not count.

The remaining wiring defects. Two criteria read an HGVS display string and broke when that string changed (v28 to v35, found as regressions), and PVS1 read the end of a deletion’s span and not the offset the span covers, which cost 14 ClinVar-pathogenic donor and acceptor deletions their PVS1 (v26, found by mining the discordant calls).

**Supplementary Note 2. Two label decisions of the scoring script.**

The VUS class is the 295,298 variants that submitters classify as uncertain without conflict, except that two RYR1 records of expert-panel review whose classification is conflicting are labelled VUS by the label script, and fastVEP calls both Likely pathogenic. Four records of CFH and CFI whose classifications are Likely pathogenic and Likely pathogenic with low penetrance are labelled Pathogenic by a pattern match of the same script. We kept both labels so that the 37 runs stay comparable: relabelling the four changes exact agreement by one variant and agreement in direction by none, and leaving out the two RYR1 records lowers the number of ClinVar VUS called pathogenic-side by two.

**Supplementary Note 3. The review rounds, and how the rulings were coded.**

She returned annotated workbooks in rounds 1, 2 and 6, with the verdict recorded as a cell colour in rounds 1 and 2 and as text in round 6, and a covering email in round 2; rounds 3 to 5 were tables and notes we sent after re-runs, which were not returned marked. The workbook of round 1 carries a list headed “Discordance categories” that names six kinds of discordance, and a key of two colours (a correct call, and both calls discordant with her own call); the key of round 2 adds a query, a request for more evidence, and a marker for genes whose population frequencies cannot be relied on because of homology. We decoded the colours against each workbook’s key, and read any green on a call as that call being correct (in round 2 the green on the cells is a different shade from the key’s, and in round 1 three greens on fastVEP’s call are darker than the key’s). In round 6 the workbook asked for her class and for who is right in two columns, which came back empty, and she wrote their ruling as free text in the verdict column of all 64 rows and added a note to 18. The round-6 text was coded by us from its content by five rules given in Appendix Table A4, and she did not assign these categories. Between rounds her notes were carried onto the rows they belong to, and each round was answered with a note that said what had changed, what it cost, and what was measured and not done. The two inputs the reviewer supplied to the classifier are ledger items 50 and 63.

What the reviewer ruled. The medical geneticist returned annotated workbooks in three rounds (Figure S3A, Appendix Table A4). In round 1 the workbook held 101 opposite-direction calls from v7, and she judged ClinVar’s call correct on 86, fastVEP’s on 3, neither on 8, and left 4 with a colour that was not in the key. Round 2 held 122 calls from v8, of which 93 were new and 29 carried over: ClinVar’s call was judged correct on 65, fastVEP’s on 2, neither on 8, more evidence was needed on 1, and 46 carried no recorded verdict. Seven of the 65 (four in CYP21A2 and three in STRC) carry the marker for a gene with a homology problem, and on six of them it is on fastVEP’s call as well as the green on ClinVar’s; the project’s note of that round counted those six as not marked, which gives 59 and 52 where we count 65 and 46. Round 6 held 64 calls from v26: by our coding of the returned text, she supported ClinVar on 43, fastVEP on 4, neither on 9, and held 8 pending more evidence or by disorder. So when the software and ClinVar disagreed in direction, the reviewer sided with ClinVar on 89% of the 97 rows of round 1 that carried a verdict in the key, 86% of the 76 rows of round 2 that carried one, and 67% of the 64 rows of round 6. The three percentages come from different instruments, which are colour decoding against a two-colour key (round 1), colour decoding against a richer key (round 2) and our coding of free text (round 6, where 14 rows were coded from the evidence they cite and not from a stated class), so they are not one series and are by our coding. The list of discordance categories that came with round 1 names six kinds of discordance: the use of PM1, PS1 or PM5 where it does not apply, a pathogenic call that rests on a founder or low-penetrance variant, and loss of function that is not the disease mechanism. Three of the kinds match what the ledger later recorded: PM1 and PS1 applied where they did not apply (items 17 and 23), a founder or low-penetrance variant that is an exception to the benign criteria (items 26, 52 and 63), and loss of function that is not the mechanism (item 33).

The first review saw about a third of the discordant calls. The workbook of round 1 listed 101 opposite-direction calls, but v7 had 313. The scorer wrote its discrepancy log with a cap of 10,000 rows; the soft discrepancies (a ClinVar pathogenic call scored VUS) filled it before the end of the genome, and the export stopped at chromosome 5, silently dropping every opposite-direction call after it. The cap was removed for opposite-direction calls in v8, whose export is complete, so 212 of v7’s 313 had never been listed. Nothing about the 101 rows was wrong, but the sample was the first five chromosomes and not a random one. It was enough to expose three gaps in how criteria were executed (PVS1’s grading, BP4’s SpliceAI path and PM2’s gnomAD lookup) and no more. Two of the seven changes that we found by reading the discordant calls ourselves, the self-matches of PS1 and PM1 and the two modes of reporting that followed, came from reproducing the reviewer’s rows, which could not have shown them.

**Supplementary Note 4. Validation of the offline replay.**

The replay was checked against real runs of the same configurations (Appendix Table A2): the Richards 2015 table on both sides, and the point cutoffs reported in the Results. It reproduces the table run, the control and 6 of the 8 runs with a changed cutoff cell for cell, and the other two (Likely pathogenic from 4 and from 5 points) within 3 and 1 variants, each a variant that the replay calls Likely pathogenic and the run, through the veto, does not. The output does not record the detail the veto needs, so the replay holds those 19 at VUS wherever it would otherwise call them Likely pathogenic.

**Supplementary Note 5. The threshold sweeps in full.**

The two PM2 thresholds. The classifier uses one PM2 threshold for genes that it does not read as recessive-only (dominant, both, or of unknown inheritance, where inheritance is read from the names of the diseases in the ClinGen file: of its 2,924 genes the classifier reads 67 as recessive-only, 50 as dominant-only, 12 as both and 2,795 as neither, see Discussion) and another for genes that it reads as recessive-only. It reaches 62.48% at the default 4e-5, and the two steps above that change it by 6 and 5 variants. Reference-discordant pathogenic calls rise from 22 at strict absence to 25 at 4e-5 and 26 at 2e-4, and agreement in direction is higher at strict absence (523,414 calls) than at the default threshold (522,886), because ClinVar-VUS variants that carry PM2 plus other evidence stay VUS. The threshold for recessive-only genes, which the code attributed to the SVI’s recommendation, which gives no frequency, and which equals the recessive threshold of the Hearing Loss expert panel, is flat as well: from strict absence to 5e-4, recall of pathogenic-side variants is between 62.46% and 62.48%, agreement in direction varies by 10 calls, and the numbers of reference-discordant benign calls and reference-discordant pathogenic calls do not change. The two thresholds therefore act on nearly every gene and on 67 genes, and a flat sweep says what a threshold does to the calls it reaches, not what a threshold would do if inheritance were read from a correct field. The sweep that v10 recorded then gave 37.8% at strict absence and 56.8% at 4e-5, against 38.8% and 56.7% here.

Recall of pathogenic-side variants under the 2015 table is 38.8% at strict absence, 46.8% at 1e-6, 54.1% at 1e-5, 56.7% at 4e-5 and 57.4% at 1e-4.

The BS1 threshold, step by step. Between 2e-2 and 5e-3 (default), lowering the threshold buys 11,489 benign-side calls at the cost of 2 reference-discordant benign calls. Below the default value the exchange rate is 928 calls per reference-discordant benign call from 5e-3 to 2e-3, 625 from 2e-3 to 1e-3, and then 112 from 1e-3 to 5e-4 and 85 from 5e-4 to 2e-4.

BP7’s intronic distance, step by step. Moving it from 200 to 300 nucleotides buys 157 correct benign-side calls for one more reference-discordant benign call, and moving it from 300 to 500 buys 40 for one more, with 51 for two more beyond that and 457 for fourteen with no bound at all. The default 300 is at the corner: below it a step is worth 157 to 849 calls per reference-discordant benign call, above it about 26 to 40.

The points scale, in detail. Variants with 0 or 1 point (27,027 and 105,989 of them) are 3.9% and 7.9% ClinVar pathogenic-side. At 2 points the share is 10.7%, at 3 points 21.9%, and at 4 points 26.5%.

The published VCEP thresholds. That median leaves out the panels that give no single number: of 137 PM2 entries the extraction took 64 as thresholds, and set aside 37 that require absence, 14 with no threshold, 9 whose threshold depends on inheritance (the Hearing Loss panel’s recessive threshold of 7e-5 among them), 6 in superscript notation, 4 where two panels disagree and 3 that are ambiguous or unspecified (Appendix Table A5). Across BA1, BS1 and PM2 together the extraction read 411 criterion entries and took 286 as thresholds (Table 1).

BP7’s PhyloP threshold, step by step. Raising the threshold from 1 to 1.5 gains 5,520 correct benign-side calls with no added reference-discordant benign call, and from 1.5 to the default 2 gains 4,571 for 3 more (1,524 per added error). Beyond the default value, raising it to 3 gains 6,539 for none, to 4 gains 3,520 for one, and switching the test off gains 6,423 for one more.

The BS2 thresholds. The steps from 1e-3 to 1e-6 are worth 463, 291 and 533 calls per reference-discordant benign call, so no setting is picked out by the data and the choice rests on how the two errors are weighed. Switching the carrier threshold off, so that an allele count of at least five alone suffices, and setting it at 3e-3 change the numbers of reference-discordant benign and reference-discordant pathogenic calls by nothing (24 and 25 throughout), on the 50 genes that the classifier reads as dominant-only. The default 3e-4 recovers 70 pathogenic-side calls relative to no threshold (58,540 to 58,610) and gives up 474 benign-side calls (177,680 to 177,206), and nothing above it recovers another.

The count of BP1’s veto, above and below the default value. At a count of 2 without the own-record change the figures are 849 more calls in direction, 23 reference-discordant benign calls (IQSEC1 removed) and 17 correct benign-side calls given up. Above the default value each step buys correct benign-side calls at the price of reference-discordant benign calls: a count of 5 gives 3 more, 10 gives 5 more and 25 gives 12 more.

The point cutoffs, step by step. A cutoff of 4 reaches 76.7% recall, but 17,452 ClinVar-VUS and 139 benign-side variants are then called pathogenic-side, and agreement in direction falls below the default value. Raising the cutoff to 7 or 8 gives up 3,257 or 3,532 pathogenic-side calls and avoids 3 reference-discordant pathogenic calls, and at 9 a lone PVS1 (8 points) is no longer Likely pathogenic, recall falls to 52.8% and reference-discordant pathogenic calls to 11. At the cutoff of 5, agreement in direction rises by 4,576 calls, and exact agreement falls by 3,624. Moving the Pathogenic cutoff between 9 and 12 changes agreement in direction by at most 3 calls, because the cutoff separates two tiers of the same direction.

**Supplementary Note 6. Where the VUS calls come from, and the baseline rules in full.**

Where the 35,178 pathogenic-side variants called VUS come from. They are not an even sample. By the evidence that was met, 10,541 (30.0%) reached 5 points, one short of Likely pathogenic, 10,597 (30.1%) reached 0 or 1 point, and 14,027 reached 2 to 4 points; 13 reached 6 points or more and were held at VUS by a veto or by benign evidence. By consequence, 18,992 (54.0%) are missense variants, 9,129 (26.0%) frameshift or nonsense, 2,041 (5.8%) in flanking or non-coding sequence, and the rest splice-region, canonical splice, in-frame, start-loss and stop-loss, intronic, UTR and synonymous variants. The concentration in genes is high: the 10 genes with most of them hold 11.7%, the top 100 hold 40.5% and the top 500 hold 74.7%, of 2,716 genes with any (CFTR 778, APC 712, LDLR 517, PAH 453, GCK 344, ABCA4 298, GLA 264, BRCA1 255, NF1 253 and SACS 246). The validity gate has one leak that changes no call: of the 505 genes whose ClinGen curations are all weak, one (NSMCE3) escapes it because its label ends in a semicolon, which the loader reads as a separator, and it has no variant in the benchmark. Absence has the effect here that it had in the first version of the validity gate (v10, Table 1), which blocked PVS1 where a gene had no curation and which v11 replaced with a gate that blocks only on curated-negative evidence.

Where the 106,937 benign-side variants called VUS come from. Of the ClinVar benign-side variants that are called VUS, 31,912 (29.8%) carry PM2 and nothing else (they are rare in gnomAD and no benign evidence fires), 20,604 (19.3%) carry BP4 with PM2, 17,467 (16.3%) a lone BP4, 14,431 (13.5%) no criterion at all, 5,586 (5.2%) a lone BS1, and the rest smaller combinations. Under the 2015 table that the benign side keeps, one supporting benign criterion does not reach Likely benign, and the Results show what the alternative would cost. At least 52,516 of these variants (49%) carry PM2, which is rarity and so does not separate a benign variant from a pathogenic one.

The 3,612 and 4,616 variants where v37 commits and ClinVar’s submitters see insufficient evidence are not obviously errors. The pathogenic-side calls are PP2 with PM2 and PP3 at Strong strength (1,195), PVS1 with PM2 (1,135), PM1 with PM2, PP2 and PP3 at Moderate strength (587), PM1 with PM2, PM5 and PP2 (448), and a lone PVS1 (163). The benign-side calls are chiefly BP1 with BP4 at Supporting or Moderate strength, with or without PM2 (3,677 of the 4,616). Each pathogenic-side call stands on 6 or more points of computable evidence, which counts correlated predictors (PP2, PP3 and PM1 for a missense variant) as if they were independent, as the point system does, and each benign-side call on two or more benign criteria or on one at Very Strong or Stand-alone strength (122 of the 4,616). A ClinVar VUS can mean that submitters weighed the evidence and found it insufficient, or that they did not weigh it to a class. The benchmark cannot say which is right, and the Results ask whether ClinVar later moved toward them.

The baseline rules in full. A higher threshold has fewer contradictions and agrees less, 52 contradictions at 59.7% for 1e-2 and 22 at 57.1% for 2e-2, so with the number of contradictions that v37 has the rule agrees for about 18 points less. The last rule is not a bargain by the standard set out in the Methods either: over the two-rule version at 1e-3 it adds 86,510 correct benign-side calls for 863 more reference-discordant benign calls, 100 per error. The population maximum used by these rules is a point estimate over ten groups, so it is more aggressive than the filtering allele frequency that BA1 tests (in v37 a point estimate in place of that frequency costs 8 more reference-discordant benign calls, see Results), but it is the threshold that sets the contradictions of the first rule (22, 52, 159 and 813 at 2e-2, 1e-2, 5e-3 and 1e-3). Of the 9,967 pathogenic-side calls that the second rule makes correctly and v37 leaves as VUS, 5,749 carry no PVS1 in v37 and 4,218 carry it at a lower strength or held back, and 43 of its 54 reference-discordant pathogenic calls are VUS in v37.

Recall by consequence class and by review status. Variants called pathogenic-side are almost all null variants: 54,498 of the 58,610 correct pathogenic-side calls (93.0%) are frameshift, nonsense or canonical splice variants, and 4,109 are missense (Figure 5A, Supplementary Table S5). Among ClinVar’s pathogenic-side variants, 88.4% of canonical splice variants and 82.6% of frameshift and nonsense variants are called pathogenic-side, against 17.8% of missense variants, 0.3% of in-frame variants and none of the start-loss, stop-loss, splice-region, intronic, synonymous, UTR and other variants. Among ClinVar’s benign-side variants the picture is the reverse: 79.2% of intronic, 65.7% of synonymous, 62.5% of UTR and 49.8% of missense variants are called benign-side (Figure 5B). The 3-star records, which are expert-panel classifications, are called pathogenic-side more often than the 2-star records (67.8% against 61.7% of their pathogenic-side variants), and called benign-side much less often (36.6% against 62.8% of their benign-side variants, with 63.3% left as VUS). Expert-panel benign-side classifications can rest on disease-specific frequency thresholds, and the published thresholds are stricter than ours (see Results), which is a possible reason for the difference and one that this benchmark cannot establish. All 23 four-star records (practice-guideline classifications) are variants of CFTR, and v37 calls 22 of them VUS (Figure 5A). Its 4,017 Pathogenic calls are ClinVar pathogenic-side for 3,996 (99.5%) and VUS for 21, and its 67,700 Benign calls are ClinVar benign-side for 67,455 (99.6%) and VUS for 245; the 25 reference-discordant pathogenic calls are all Likely pathogenic and the 24 reference-discordant benign calls all Likely benign.

**Supplementary Note 7. The rest of the PVS1 gap.**

Of the 5,184 truncating variants that carry no PVS1, 625 genes hold them (620 by the symbol of the annotation, which differs from ClinVar’s first gene symbol for 55 of the variants). The second route to loss of function is nearly empty: of the 26 genes in the default table of mechanisms, 20 are gain of function, one is dominant negative and five name both loss and gain of function, and those five are each licensed by constraint or by a curation already. All 76 variants in genes that gnomAD constrains (RASA1 40, TGFB3 13, LRP2 11, VPS13D 5, SLC12A5 3, DMXL2 2 and PTPN11 2) are held by a gate, 74 by the validity gate and 2 by the mechanism gate (Appendix Table A8). The only curation of LRP2 is “no known disease relationship” with congenital heart disease, where ClinVar’s records name Donnai-Barrow syndrome for 10 of its 11. The ClinGen guidance on genes with several disorders (ClinGen, 2024), whose per-disorder scaffold is in the classifier and is not active, is the answer to that. The other 3,945 truncating variants that are called VUS carry PVS1 at a lower strength (3,062 at Strong, 463 at Moderate, 416 at Supporting and 4 at full strength but held back by other evidence), and 1,434 of the 1,446 canonical splice variants that are called VUS carry no PVS1 either.

The completed ClinGen file in detail. The completed file adds the 112 rows of the October export that are dated on or before 30 April and whose gene and disease pair the April file lacks: the 104 curations, seven rows that re-label, under another disease identifier, an assertion that the April file lists, and one row (GATM, Moderate) dated 04:00 UTC on 30 April, which is after the April file was saved (02:19 UTC, by its modification time) and changes none of the 163 calls on GATM’s variants. The 674 variants that gain PVS1 are in 23 genes: CFTR 439, ELP1 85, EVC 46 and SPG7 41 among them, which are 611 of the 674. The two reference-discordant pathogenic calls the run adds are both frameshift variants of ALPK1 that ClinVar calls Benign or Likely benign at allele frequencies of 2.3e-4 and 6.4e-4. Orphanet’s label is the association type “disease-causing germline mutation(s) (loss of function)” of Orphadata product 6, version of 23 June 2026, where a gene that also has a gain-of-function association is taken as both (Orphanet, 2026), and the 5 reference-discordant pathogenic calls it adds are MAP3K6, ALPK1 twice, VPS13C and LTBP4, two of them splice-acceptor variants with population-maximum frequencies of 0.8%.

**Supplementary Note 8. The residual missense calls, and where the rulings come from.**

Missense variants: PM1, PP2 and PP3 on a variant ClinVar calls benign or likely benign (3), BS1 on pathogenic missense variants (2) and BP1 on pathogenic missense variants (2). The first three (CHD7, FBN1 and PTCH1) reach 6 points through PM1, PM2, PP2 and PP3 at Moderate strength, and she supported ClinVar on all three. The BS1 pair (PADI3 and SLC12A3) are seen in 328 and 338 gnomAD alleles, with population-maximum frequencies of 0.6% and 0.8%; she noted a homozygote in gnomAD and a disorder (Gitelman syndrome) for SLC12A3, supported ClinVar there, and asked for a deeper look at PADI3. The BP1 pair (IQSEC1 and PRR12) are in genes with two and one pathogenic or likely pathogenic missense records in the ClinVar protein index, the variant’s own among them, so BP1’s veto does not apply (a count of 1 would withhold BP1 for both, for PRR12 only through its own record and for IQSEC1 also through p.Thr343Met, see Results), and she supported ClinVar on both. Of the 11 variants on which PVS1 fired alone, and the 11 on which it fired with PM2, all are loss-of-function variants in a gene that the classifier takes to be sensitive to loss of function. Of the 30 rulings for ClinVar, 11 follow from a class that the reviewer stated, 9 from the reviewer’s statement that ClinVar is correct, and 10 from evidence that the reviewer gave for ClinVar’s class without naming one (Appendix Table A4). Of the 15 calls that were repaired between v26 and v37, she had supported ClinVar’s call on 13; of the other two, one was coded neither and one depends on the disorder. One of the 20 BP4 and BP7 calls is POLR3A c.1909+22G>A, at an allele frequency of 2.5e-3 in gnomAD, for which she cites affected individuals and some functional data.

**Supplementary Note 9. The eight departures from the sources, in full.**

First, PVS1 grades an NMD-escaping truncation outside a critical region as Moderate when at least 10% of the protein is removed and Supporting when less is, where the tree of Abou Tayoun et al. (2018) gives Strong and Moderate, and it grades a canonical splice variant in the last exon as Moderate whatever the fraction removed, where the tree has no last-exon branch and grades by critical region and fraction. Regrading both by one step would change 7 of 673,660 calls (6 from VUS to Likely pathogenic and 1 from Benign to VUS), by a replay that is an upper bound, because the recorded code does not say which branch gave a PVS1_Moderate; so few change because a grade one step lower plus PM2 seldom reaches 6 points either way. Second, PVS1 predicts NMD by whether the variant lies in the last exon, where the tree predicts it by the 50-nucleotide rule, which also lets a stop in the last 50 nucleotides of the penultimate exon escape; the exact rule is a setting, the code comment calls it the guideline-faithful one, and it is off by default because of what it costs against ClinVar: 603 calls in direction and 0.73 points of pathogenic-side recall, to avoid one reference-discordant pathogenic call (see Results). That is a default chosen on agreement with ClinVar, a tuning to the reference that is not the dependence on ClinVar-derived inputs that the Results measure. Third, BP7 keeps the 2015 test of poor conservation (PhyloP below 2.0) and takes a SpliceAI score below 0.2 as no impact, where Walker et al. (2023) apply BP7 after BP4, which for SpliceAI is a score of 0.1 or below, and caution against a conservation test. Fourth, the allele-number floor has a narrower source than its use: BA1 and BS1 require an allele number of at least 2,000 at the variant’s site in gnomAD, a figure that comes from the BA1 recommendation, which asks for an allele frequency above 5% in a general continental population dataset of at least 2,000 observed alleles (Ghosh et al., 2018), whereas the project’s documentation attributed it to the March 2024 communication on gnomAD v4, which states no allele-number floor and recommends the grpmax filtering allele frequency (ClinGen VCEP Review Committee, 2024); here it is applied to the allele number at the variant’s own site, and to BS1 as well, and neither use is stated by either source. Fifth, the recessive-gene PM2 threshold of 7e-5 was attributed in the code to the SVI’s PM2 recommendation, which gives no frequency at all; the number equals the recessive threshold of the Hearing Loss expert panel, and the project’s record does not say where it came from. Sixth, the veto of BP1 counts the variant’s own ClinVar record, which PS1 and PM1 exclude. Seventh, the ClinGen file that the benchmark read lacked 104 curations that ClinGen dates before the day of the export, among them CFTR. Eighth, three citations were wrong: the PMID of Ghosh et al. (2018), which was wrong in three places, the journal of Tavtigian et al. (2020), which is Human Mutation and not Genetics in Medicine, and the date of the ClinGen guidance on genes with multiple disorders, which is January 2024 and was recorded as July 2025. We corrected the citations and the comments in the repository. Two comments in the classifier were wrong and are corrected: the one that said the LOF_and_GOF mechanisms are inert, which in fact license PVS1 through the curated-mechanism route and change no call in the benchmark, and the one that said the ClinGen phenotypes are Moderate or better only, where they are every classification and the validity gate is what withholds the weak ones.

**Supplementary Note 10. The opt-in settings, and the two trades in the other direction.**

Two opt-in settings change little: capping PP3 at Moderate raises agreement by 712 calls and lowers pathogenic-side recall by 0.34 points with no change in errors, and the exact 50-nucleotide NMD rule for PVS1 loses 603 calls to avoid one reference-discordant pathogenic call. Two settings would trade errors for recall in the other direction. PM2 at Moderate, as in the 2015 standards, raises recall of pathogenic-side variants by 11.0 points (10,280 more variants) and agreement in direction by 4,475 calls, for 14 more reference-discordant pathogenic calls (about 320 calls per error). The 2015 table on both sides loses 5,255 calls and 5.8 points of recall to avoid 11 reference-discordant pathogenic calls, which is the combining-rule effect reported in the Results. Neither is a clear improvement by the standard of this paper, and the first contradicts the ClinGen recommendation that PM2 be used at Supporting strength.

**Supplementary Note 11. The new Benign variants that frequency did not reach.**

What the mix of frequency bands does not explain is mostly in the highest bands (the two at 2% or more carry 1.8 of the 2.7 points that the mix does not explain): the new Benign variants at 2% or more are called benign-side for 58% (326 of 564) against 91% of 83,527 April ones, and 157 of the 238 that are not carry BP4 alone, so neither BA1 nor BS1 fired on them. Annotating those 238 again with the details of each criterion shows why. For 189 (79%) BA1 and BS1 were withheld by design: 124 sit in a low-complexity region that gnomAD flags, 56 at a site whose gnomAD record fails its own filter, 5 in a segmental duplication and 4 in a gene of the homologous-gene list, each counted once, by the first of these that applies (see Results). For 17 (7%) the criteria were evaluated and the filtering allele frequency is below the threshold, which is how one small group can inflate a population maximum. Another 31 met BS1 and stay at VUS, 23 with BS1 alone, which the 2015 table on the benign side does not call, and 8 with pathogenic-side evidence beside it. One more has an allele number below the floor. The 189 are 34% of the 564 new Benign variants of that frequency, against 4.8% (4,035 of 83,527) of the April ones, so the new variants are far more often at sites where the classifier does not trust the gnomAD frequency, and we have not established why. Recall of ClinVar Pathogenic variants is 64.7% against 65.0%, and of Likely pathogenic variants 51.4% against 48.3%. We made the split into nine bands after a split into six, with one band from 1e-2 up and one from 1e-3 to 1e-2, which explained 70%, had shown where the rest lay, so the 86% describes the fall and does not test it. The 18 opposite-direction calls on the new variants are, by gene: 9 PVS1 calls on ClinVar benign-side variants (ALMS1, CACNA1G, CCNQ, MAP3K14, ORAI1, PRKDC, SMARCA2, TRPV4, UNC93B1), 6 BP4 and BP7 calls on ClinVar pathogenic-side variants (ABCA4, HEXA, IFT122, NUP107, PKD1, SLC7A9), and 3 missense variants called pathogenic-side on PP2 and PP3 (COL1A1 twice and KCNH1).

**Supplementary Note 12. What the later release does not test.**

The new variants lie in 4,124 genes, all but one of which are in the April set, so this is a hold-out on variants and not on genes, and every gene-level setting (the exception list, the homologous-gene list, the ClinGen gene tags) was set with the April genes in view. The ClinVar-derived indices the classifier reads were not rebuilt, so the protein index lacks every record added since April, and holds the April record of each new variant that was then a classified missense variant, which for BP1’s veto is the variant’s own record (for 77 of the 94 new ClinVar-pathogenic missense variants in genes where BP1 can fire, and for the other 17 the April index does not hold it as pathogenic), as it is for 788 of the 790 April variants of the same kind. The records of other variants that were added or reclassified since lower PS1 and PM5 for a new variant, lower or raise PM1 (fewer pathogenic neighbours lower it and fewer benign ones raise it), and make the veto of BP1 less likely to apply, which could add reference-discordant benign calls. A new variant that already has a pathogenic record in the allele-level annotation of 27 June has that record left out of the index counts of PS1 and PM1 although the April index does not hold it, which lowers both further. None of the 18 opposite-direction calls on the new variants rests on BP1, PS1 or PM5 and one rests on PM1, so the missing records can have produced at most one of them. And six months is short: ClinVar reclassifies slowly, so the later release adds mostly second submissions to variants that ClinVar already held, and little new knowledge about old variants. The measurement build discounts one record whenever ClinVar’s allele-level record of 27 June is pathogenic, which for up to 17 of the 94 (those whose own record the April index does not hold as pathogenic) discounts a record that is not in their count, and the run changes no call for them either.

#### Supplementary tables

A ClinVar class in these tables is the class of a ClinVar 2-star+ record of the release of 4 April 2026, and of 4 October 2026 where a table says that it is about the later release.

**Supplementary Table S1.** The 28 criteria and the benchmark: what each needs, whether the benchmark can apply it and, on every 40th record of the benchmark annotated to JSON, how often the engine could evaluate it and how often it met it (criteria_status.csv, made by 18_criteria_status.py); and how often each of the 15 computable criteria fires in v37, at which strengths, and on variants of which ClinVar class (criteria_firing_v37.csv).

**Supplementary Table S2.** The versions and the runs of this paper. For 36 of the 37 versions (v3 was a partial run that was overwritten before it was preserved), the agreement, the opposite-direction calls and the recall of each, with the number of variants each committed to, the denominator of each, and the marker of v17 (trajectory.csv; v1 is a reconstructed matrix and v2 and v4 to v6 are the figures recorded at the time). For v37 itself, the 89 runs made for this paper and the 8 earlier runs of the dominant-gene BS2 settings (98 rows), the concordance matrix, the firing counts and the settings (runs_v37_variants.csv; the run matrix, the matrices of the seven runs on the October release and the provenance files are in analysis/acmg_benchmark/results/v37_manuscript/).

**Supplementary Table S3.** The ledger of 64 changes: run, class, route of discovery and source in the project record (change_ledger.csv).

**Supplementary Table S4.** What each step of each threshold sweep buys and costs (threshold_steps_v37.csv), and the median, quartiles and range of the published VCEP thresholds (vcep_bars_v37.csv).

**Supplementary Table S5.** Concordance of v37 with ClinVar 2-star+ classifications: the five-tier and three-tier matrices, what ClinVar says behind each call, the concordance by consequence class and by review status, the 23 four-star records, and the headline rates with their intervals (confusion_5x5_v37.csv, confusion_3x3_v37.csv, what_is_behind_each_call_v37.csv, by_consequence_v37.csv, by_review_status_v37.csv, four_star_variants_v37.csv, headline_v37.csv).

**Supplementary Table S6.** The rules with no combining rule, gene-level gate or safeguard and what they agree with ClinVar on, at one threshold and at nine, beside v37; the second rule against the PVS1 gap; and what each tier of the v37 call is worth, with the rate of opposite-direction calls and its interval (baselines_v37.csv, baselines_curve_v37.csv, baselines_r2_vs_v37.csv, precision_by_tier_v37.csv).

**Supplementary Table S7.** The PVS1 gap and the ClinGen file: truncating variants without PVS1, by gene and by the reason PVS1 is absent (pvs1_gap_v37.csv, pvs1_gap_genes_v37.csv, pvs1_gap_reasons_v37.csv); the 112 rows that complete the ClinGen export of 30 April, 104 of them curations that it lacked (clingen_missing_curations.csv, clingen_missing_summary_v37.csv); the 558 null variants of CFTR, each with its transcript, its call in v37 and the call that the completed file gives (cftr_null_variants_v37.csv); and what the three runs that change an input do to the gap (pvs1_gap_closure_v37.csv).

**Supplementary Table S8.** The 49 opposite-direction calls of v37 with their criteria, gnomAD counts and rulings, and the rulings by kind of call (opposite_direction_v37.csv, opposite_direction_by_group_v37.csv).

**Supplementary Table S9.** The later ClinVar release: the strata, recall by class, coverage, the opposite-direction rates and their ratio with its interval, the 18 opposite-direction calls on the new variants, and the seven proposed changes run on the new variants (temporal_v37.csv, temporal_by_class_v37.csv, temporal_coverage_v37.csv, temporal_opposite_rates_v37.csv, temporal_ratio_v37.csv, temporal_new_opposite_v37.csv, temporal_proposals_v37.csv).
